# Multiple imputation of missing data under missing at random: compatible imputation models are not sufficient to avoid bias

**DOI:** 10.1101/2022.11.04.22281883

**Authors:** Elinor Curnow, James R Carpenter, Jon E Heron, Rosie P Cornish, Stefan Rach, Vanessa Didelez, Malte Langeheine, Kate Tilling

**Affiliations:** Department of Population Health Sciences, Bristol Medical School, University of Bristol, Bristol, UK; Medical Research Council Integrative Epidemiology Unit at the University of Bristol, University of Bristol, Bristol, UK; Department of Medical Statistics, London School of Hygiene and Tropical Medicine, University of London, London, UK; Medical Research Council Clinical Trials Unit at University College London, University of London, London, UK; Leibniz Institute for Prevention Research and Epidemiology - BIPS, Bremen, Germany; Faculty of Mathematics/Computer Science, University of Bremen, Bremen, Germany; Senator for Health, Women and Consumer Protection, Bremen, Germany

## Abstract

**Background:** Epidemiological studies often have missing data. Multiple imputation (MI) is a commonly-used strategy for such studies. MI guidelines for structuring the imputation model have focused on compatibility with the analysis model, but not on the need for the (compatible) imputation model(s) to be correctly specified. Standard (default) MI procedures use simple linear functions. We examine the bias this causes and performance of methods to identify problematic imputation models, providing practical guidance for researchers.

**Methods:** By simulation and real data analysis, we investigated how imputation model mis-specification affected MI performance, comparing results with complete records analysis (CRA). We considered scenarios in which imputation model mis-specification occurred because (i) the analysis model was mis-specified, or (ii) the relationship between exposure and confounder was mis-specified.

**Results:** Mis-specification of the relationship between outcome and exposure, or between exposure and confounder in the imputation model for the exposure, could result in substantial bias in CRA and MI estimates (in addition to any bias in the full-data estimate due to analysis model mis-specification). MI by predictive mean matching could mitigate for model mis-specification. Model mis-specification tests were effective in identifying mis-specified relationships. These could be easily applied in any setting in which CRA was, in principle, valid and data were missing at random (MAR).

**Conclusion:** When using MI methods that assume data are MAR, compatibility between the analysis and imputation models is necessary, but is not sufficient to avoid bias. We propose an easy-to-follow, step-by-step procedure for identifying and correcting mis-specification of imputation models.

## 1. Introduction

Missing data are ubiquitous in health and social research. While there are a number of methods for analysing partially observed datasets (including inverse probability weighting and maximum likelihood methods such as the expectation-maximisation algorithm), multiple imputation (MI) is the most flexible, general, and commonly used [1]. When imputation models are appropriately specified, MI gives valid inferences if data are missing completely at random (MCAR) or missing at random (MAR), but not (unless additional information is provided by the analyst) if data are missing not at random (MNAR) (Table 1). Appropriate specification of the imputation model for each partially observed variable means that (a) it must be compatible with the analysis model (*i.e*. it must contain the same variables in the same form, including any interaction terms implied by the analysis model) [2-5], and (b) it must be a correctly specified model for the variable being imputed. For example, in clinical trials the pre-specified analysis model might specify a linear time trend, but if the trial outcome data have in truth a non-linear relationship with baseline data – and this is not included in the imputation model – then we say the imputation model is compatible (with the analysis model) but mis-specified. Guidelines [1, 6] have tended to focus on compatibility rather than correct specification.

**Table 1.**
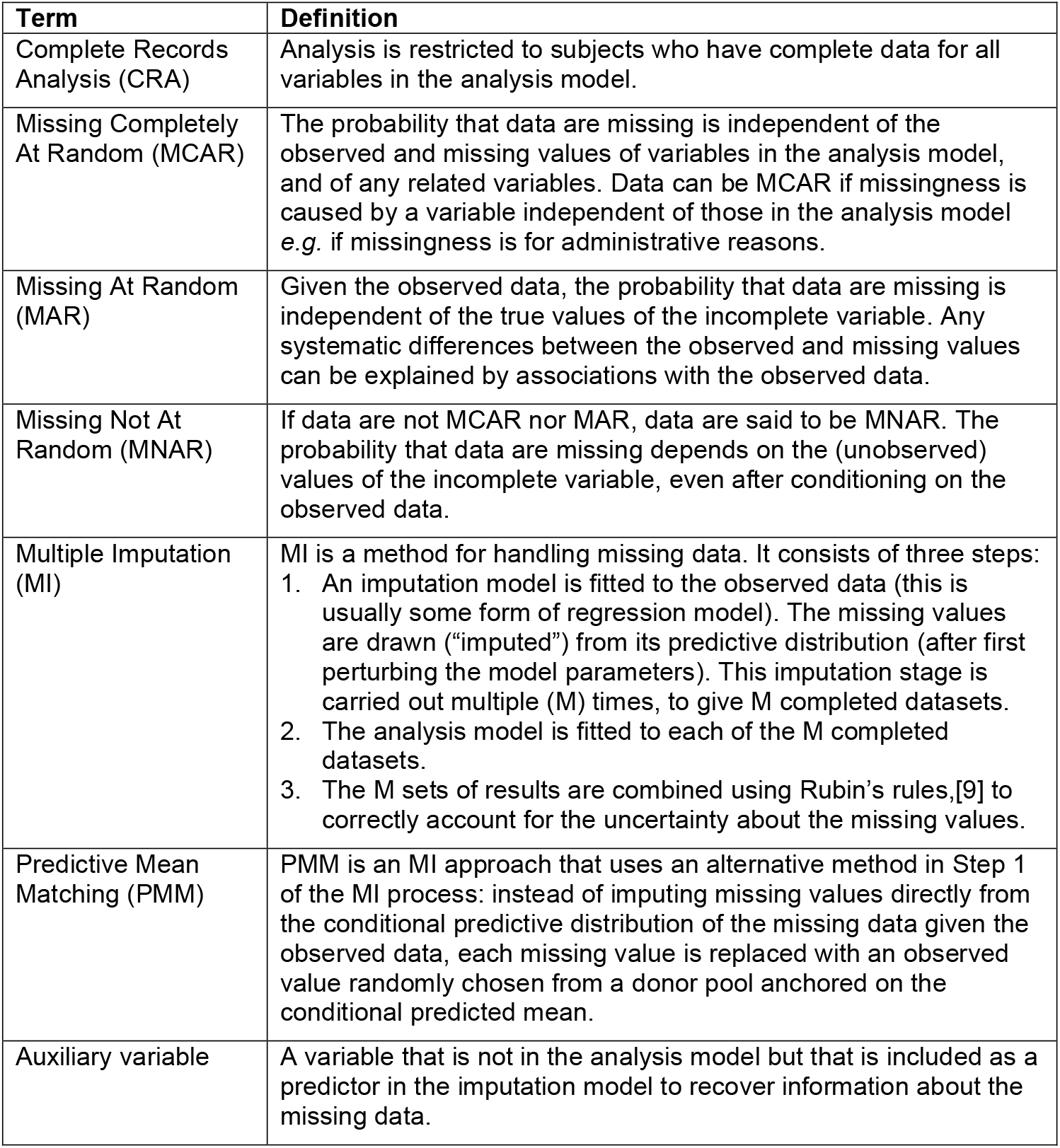
Missing data definitions

In this paper, we aim to examine the bias caused by mis-specified imputation models and provide practical guidance to researchers for identifying and correcting any imputation model mis-specification. We were motivated by the analysis of a randomised controlled trial assessing the effect of a “use acupuncture” treatment policy for chronic headache [7, 8]. The primary outcome (headache score at one year follow-up) was missing for 25% of participants. The primary analysis was a complete records analysis (CRA), and a sensitivity analysis used MI. Both analyses assumed linear relationships between the primary outcome and the continuous covariates (baseline headache score, age, and chronicity). However, an exploratory analysis described in the original publication suggested a non-linear relationship between baseline headache score and the primary outcome, which was not accounted for in the CRA or MI analyses.

Here, through a comprehensive set of simulation studies and a re-analysis of the trial data, we investigate the likely bias in CRA and MI estimates due to model mis-specification. As part of this, we compare methods (which make use of the complete records) for identifying mis-specification of imputation models. We conclude with an easy-to-follow, step-by-step procedure for identifying and correcting any imputation model mis-specification when performing MI.

## 2. Simulation study

We used simulation to assess the performance of MI when the imputation model was mis-specified, comparing the results with CRA. As mentioned earlier, we considered scenarios in which imputation model mis-specification occurred because either (i) the chosen analysis model, or (ii) the relationship between the exposure and confounder, was mis-specified with respect to the data generating model. We considered various scenarios, likely to occur in real data, in which there were non-linear relationships between an outcome *Y* (either continuous or binary), a single continuous exposure *X*, and a single confounder *C* (either continuous or binary).

The aims were (a) to quantify the bias of, and accuracy of inference for, the exposure coefficient (*β*_*X*_) and (b) to quantify the sensitivity and practical utility of available tests of imputation model mis-specification. In each scenario, the true value of *β*_*X*_ was defined as the full data estimate using all 1000 simulated datasets combined. Hence, here, “bias” refers to the difference between the CRA or MI estimate and the full data estimate *i.e*. we aimed to evaluate specifically how much bias occurred due to our analysis approach, *in addition* to any bias due to using a mis-specified analysis model. We used 1000 simulations in each scenario, and each simulated dataset contained 1000 subjects. The standard deviation of the per-simulation estimates of 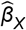 was at most 0.2 (see Supplementary Material, Tables S1-S4). Hence, 1000 simulations gave a Monte Carlo (MC) standard error [11] of the estimated bias of 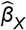 of at most 0.006. All analyses were conducted using Stata (17.0, StataCorp LLC, College Station, TX). Stata code to perform the simulation study is included in Supplementary Material, Section S11.

### 2.1 Methods for simulating full and missing data

#### Generating full data

We considered four different scenarios (Table 2), allowing for non-linear relationships between *Y* and *X* (Scenarios 1 and 4, where *Y* is continuous in Scenario 1 and binary in Scenario 4), *Y* and *C* (Scenario 2), and *X* and *C* (Scenario 3). In Scenario 1, we varied the strength of the non-linear association between *Y* and *X*. In all other scenarios, we used fixed values for non-linear associations.

**Table 2.** Data generating mechanism (DGM) and missingness mechanism used in the simulation study in Scenarios 1 to 4

| Scen. | DGM for Y | DGM for X | DGM for C | Partially observed variables* | Missingness mechanism** |
| --- | --- | --- | --- | --- | --- |
| 1 | Y is continuous and depends on X, $X^2$ and C:<br>$Y = -0.4 + 0.4 X + 0.8 C + \varphi X^2 + \varepsilon_Y$<br>where $\varphi = 0.1, 0.6$ or $1.0$ | X is continuous and depends on C:<br>$X = C + \varepsilon_X$ | C is binary, with probability 0.5 of a value of 0 or 1 | C or Y | Missingness depends on X:<br>$\text{logit}\{P(R_{\Delta}=1)\} = \tau (\alpha + X)$ |
| 2 | Y is continuous and depends on X, C and $C^2$:<br>$Y = -0.4 + 0.4 X + 0.8 C + 0.6 C^2 + \varepsilon_Y$ | X is continuous and depends on C:<br>$X = C + \varepsilon_X$ | C is normally distributed, with mean 0.5, variance 1 | X or Y | Missingness depends on C:<br>$\text{logit}\{P(R_{\Delta}=1)\} = \tau (\alpha + C)$ |
| 3 | Y is continuous and depends on X and C:<br>$Y = -0.4 + 0.4 X + 0.8 C + \varepsilon_Y$ | X is continuous and depends on $C^2$:<br>$X = C^2 + \varepsilon_X$ | C is normally distributed, with mean 0.5, variance 1 | X or Y | Missingness depends on C:<br>$\text{logit}\{P(R_{\Delta}=1)\} = \tau (\alpha + C)$ |
| 4 | Y is binary and depends on X, $X^2$ and C:<br>$\text{logit}\{P(Y=1)\} = -0.4 + 0.4 X + 0.8 C + 0.5 X^2$ | X is continuous and depends on C:<br>$X = C + \varepsilon_X$ | C is binary, with probability 0.5 of a value of 0 or 1 | C or Y | Missingness depends on X:<br>$\text{logit}\{P(R_{\Delta}=1)\} = \tau (\alpha + X)$ |

*logit*, logistic function
In each scenario, we assumed the error terms $\varepsilon_Y$ and $\varepsilon_X$ were uncorrelated, with standard normal distributions (mean 0, variance 1).
\* Values were set to missing for one variable only in each setting.
\*\* $P(R_{\Delta}=1)$ denotes the probability that a value (of the partially observed variable) is observed, with $\Delta = X, C$ or $Y$ .
For each missingness mechanism, $\alpha$ was chosen empirically to give approximately 70% observed values, for each strength of missingness association ( $\tau$ ), $\tau = 0.1, 1, 3$ or $5$ .

#### Generating missing data

In each scenario, only one variable had missing values (*i.e*. for one of *X, C* or *Y* in each setting), using MAR mechanisms (Table 2). In each scenario, we considered four different strengths of the missingness association (see Supplementary Material, Section S10, for plots illustrating the impact of the strength of the missingness association on the distribution of missing values). The percentage of cases with missing values was approximately 30% in each scenario.

### 2.2. Analysis approaches

In our analysis models, we assumed a linear relationship between *Y* (or *logit*{P(*Y*=1)} when *Y* was binary – note that for brevity, we will simply refer to *Y* hereafter), *X*, and *C, i.e*. we fitted the model E(*Y*) = *β*_*0*_ + *β*_*X*_ *X* + *β*_*C*_ *C* when *Y* was continuous, and *logit*{P(*Y*=1)} = *β*_*0*_ + *β*_*X*_ *X* + *β*_*C*_ *C* when *Y* was binary.

In the presence of missing data (for either *Y, X*, or *C*), we compared three analysis approaches:

i. CRA.
ii. MI using standard methods (imputation using a linear or logistic regression model, for continuous or binary variables respectively, hereafter referred to as MI).
iii. MI by “type 1” predictive mean matching [10] (hereafter referred to as PMM). This method was used for continuous variables only.

Following current guidelines [6], we performed MI and PMM compatible with the analysis model, *i.e*. we assumed linear relationships between *Y, X*, and *C* in the imputation model(s). We used a donor pool of size five for PMM and performed 30 imputations for both MI and PMM.

### 2.3. Tests for model mis-specification

In each setting, we tested both the analysis and imputation models for mis-specification, using the appropriate test for the model being assessed. Therefore, we considered linear regression model mis-specification tests when continuous variables were partially observed and/or the outcome of the analysis model was continuous. We considered logistic regression model mis-specification tests when binary variables were partially observed and/or the outcome of the analysis model was binary. We assessed model mis-specification by fitting the chosen model using the complete records.

Applying this approach to Scenario 1, for example, we (i) fitted a linear regression model to examine the analysis model specification, when either *Y* or *C* was partially observed, and (ii) we fitted a logistic regression model to examine the imputation model specification when *C* was partially observed. Note that, after examining the specification of the analysis model, it was not necessary to perform additional tests for imputation model mis-specification when *Y* was partially observed. This was because relationships between *Y, X*, and *C* were the same in the analysis and imputation models in our simulations. In other applications, therefore, these tests would only additionally be needed for the imputation model when it included auxiliary variables (Table 1).

In Scenario 1, we considered nine tests of model mis-specification (six for linear regression and three for logistic regression), listed below (see Supplementary Material, Section S8, for further details).

#### Linear regression model mis-specification tests

1. Pregibon [12] (link) test
2. Shapiro-Wilk [13] test
3. Breusch–Pagan/Cook–Weisberg [14, 15] (heteroskedasticity) test
4. Fractional polynomial (FP) degree-two residuals test
5. FP degree-one residuals test
6. Grouped residuals test

#### Logistic regression model diagnostic tests

7 Pregibon [12] (link) test
8 Hinkley [16] test
9 Hosmer-Lemeshow [17] test

In Scenarios 2-4, we applied one test for linear regression models and one test for logistic regression models. We chose the tests that had performed the best (in terms of test sensitivity, see Results section) in Scenario 1. We assumed that test performance would be similar in Scenarios 2-4 because the form and strength of the relationships between *Y, X*, and *C*, as well as the missingness mechanisms, were similar to those in Scenario 1.

## 3. Results

### 3.1. Results for Scenario 1: estimating the exposure coefficient

Results for Scenario 1 (quadratic relationship between continuous variables *Y* and *X*) are summarised in Figure 1. We show results using the three analysis approaches (CRA, MI and PMM) when the analysis model (incorrectly) assumes linear relationships between *Y, X*, and *C*, and missingness of *C* or *Y* depends on (fully observed) exposure *X*. We present standardised bias of 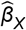, defined as 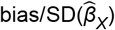, plotted against the strength of the missingness association. Full results showing standardised bias, bias of 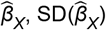, and model-based standard error are included in the Supplementary Material (Table S1). Hereafter, for brevity, we will refer to standardised bias as “bias”.

**Figure 1.**
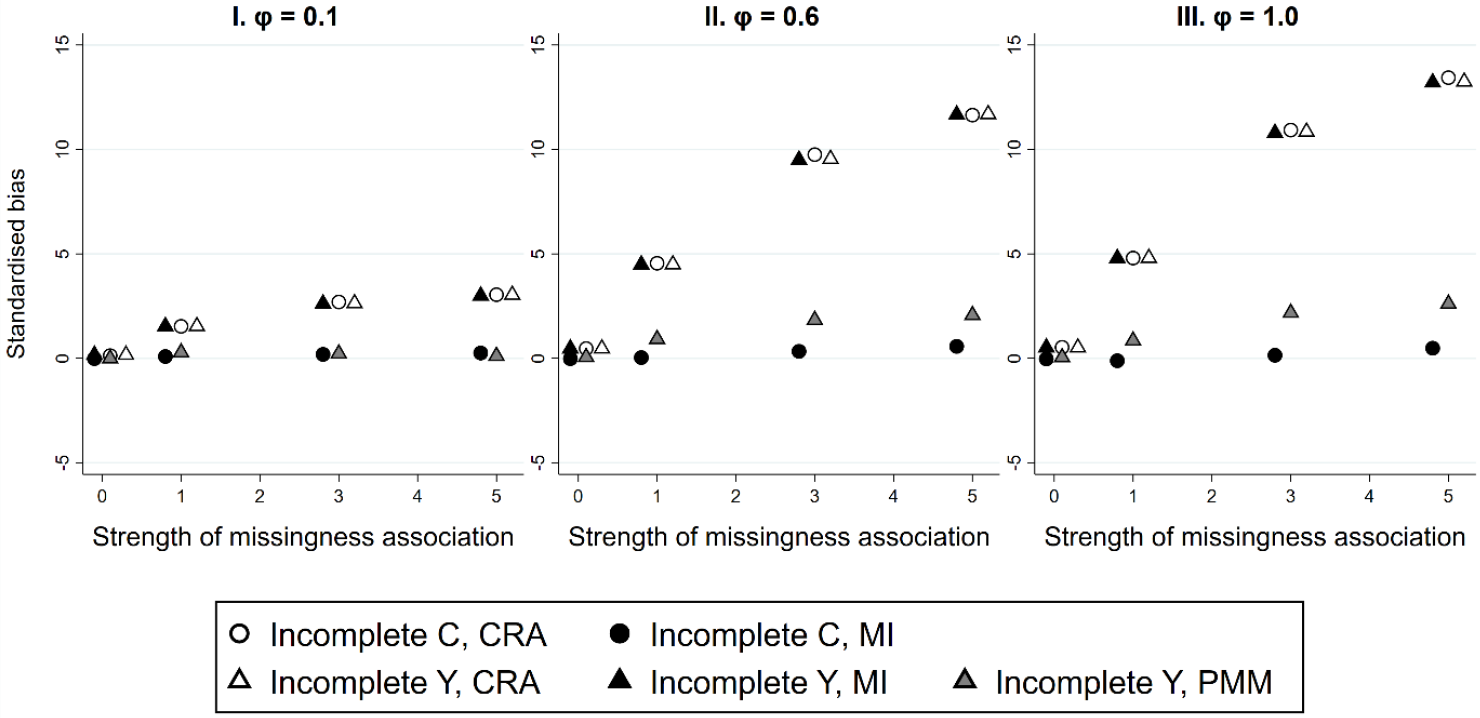
Results for Scenario 1: Standardised bias of complete records analysis (CRA), standard multiple imputation (MI), and predictive mean matching (PMM) estimates of parameter β_X_, plotted against the strength of the missingness association, for different strengths of the non-linear association (φ) between X and Y. Some overlapping points have been horizontally jittered.

In Scenario 1, neither the chosen analysis model nor the imputation model for *Y* specify the relationship between *X* and *Y* correctly. Hence, as expected, both CRA estimates when *C* or *Y* are partially observed, and MI estimates when *Y* is partially observed, are biased. Figure 1 shows that bias is of similar magnitude in each case and increases with the strength of the non-linear association (increasing across plots I-III), as well as with the strength of the missingness association (shown on the x-axis in each plot). PMM estimates are less biased than MI estimates when *Y* is partially observed, although some bias remains unless the non-linear association is weak. Conversely, MI estimates when *C* is partially observed have little or no bias, even though the imputation model is mis-specified (mis-specification occurs in this case because the imputation model for *C* should include all relationships implied by the correct analysis model, in the same form as in the correct analysis model).

### 3.2. Results for Scenarios 2-4: estimating the exposure coefficient

Results for Scenarios 2-4 (quadratic relationship between continuous variables *Y* and *C*, quadratic relationship between continuous variables *X* and *C*, and quadratic relationship between binary *Y* and continuous *X*, respectively) are illustrated in Figure 2. As before, we present standardised bias of 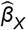, plotted against the strength of the missingness association (see Supplementary Material, Tables S2-S4, for full results).

**Figure 2.**
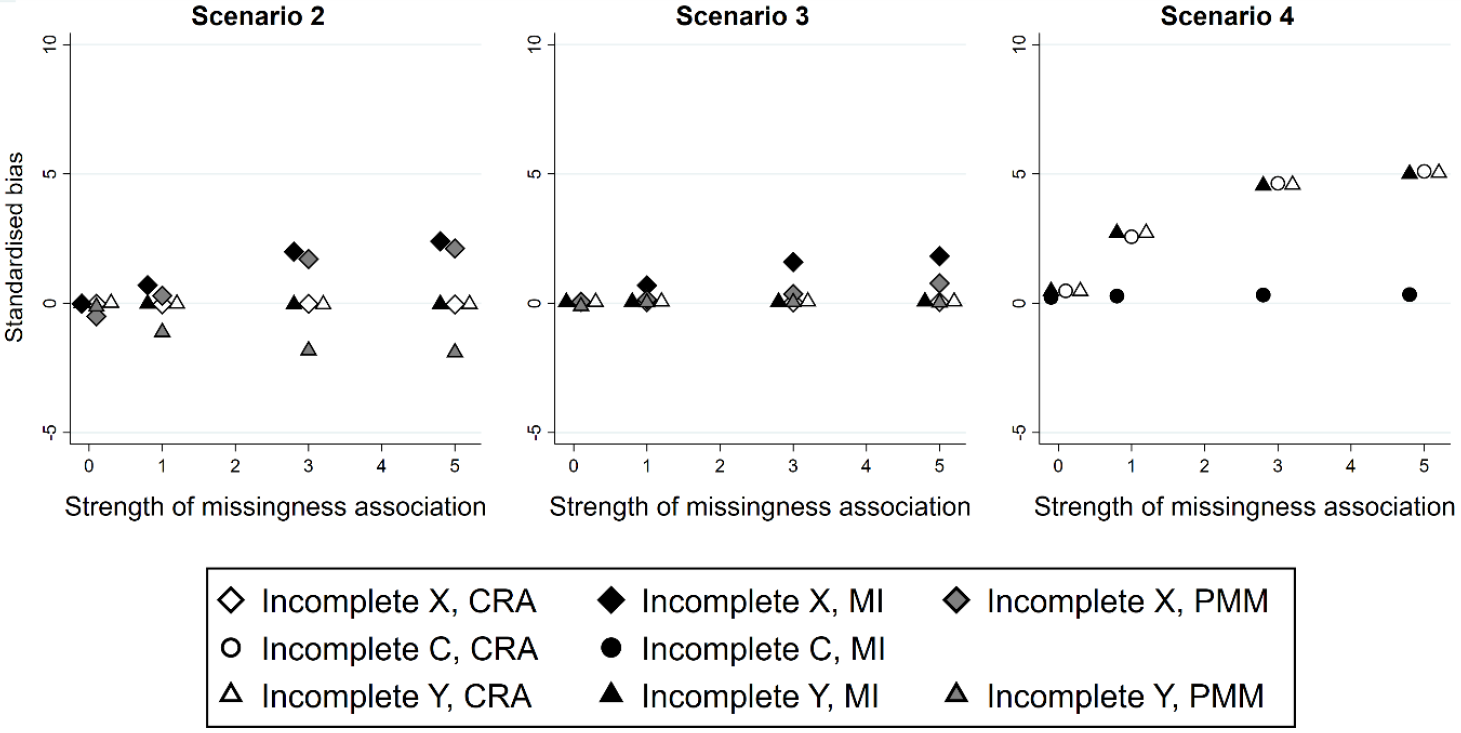
Results for Scenarios 2-4: Standardised bias of complete records analysis (CRA), standard multiple imputation (MI), and predictive mean matching (PMM) estimates of parameter β_X_, plotted against the strength of the missingness association. Some overlapping points have been horizontally jittered.

As in Scenario 1, Figure 2 shows that any bias in 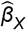 increases in magnitude as the strength of the missingness association increases. In Scenario 2, although the relationship between *Y* and *C* is mis-specified, the relationship between *Y* and *X* is correctly specified. Hence, CRA estimates of *β*_*X*_, and the MI estimate when *Y* is partially observed, have little or no bias. However, when *X* is partially observed, mis-specification of the imputation model for *X* results in a biased MI estimate (as before, mis-specification occurs in this case because the imputation model for *X* should include all relationships implied by the correct analysis model, in the same form as in the correct analysis model). In contrast to Scenario 1, the PMM estimate is more biased than CRA or MI estimates when Y is partially observed, and only slightly improves bias when *X* is partially observed.

In Scenario 3, the relationship between *Y, X* and *C* is correctly specified in the analysis model, and in the imputation model for *Y*. Hence, CRA estimates of *β*_*X*_, and the MI estimate when *Y* is partially observed, are unbiased as expected. PMM estimates when *X* or *Y* are partially observed have little bias. MI estimates when *X* is partially observed have some bias, because the relationship between *X* and *C* is mis-specified in the imputation model for *X*. In Scenario 4, results are very similar to Scenario 1.

### 3.3. Results: tests for model mis-specification

For each test of model mis-specification that we considered in Scenario 1, Tables 3 and 4 show (for linear and logistic regression model tests, respectively) the proportion of test p-values < 0.05 when the relevant model was mis-specified (*i.e*. when it assumed linear relationships between *Y, X*, and *C*), and the proportion of test p-values < 0.05 when the model was correct (*i.e*. when it additionally included *X*^*2*^). We refer to these proportions as “sensitivity” and “type 1 error”, respectively. Results are shown when testing the analysis model with *Y* partially observed (Table 3) and when testing the imputation model for *C* with *C* partially observed (Table 4), for a single value of the strength of the missingness association (*τ* = 1), and three different strengths of non-linear association between *X* and *Y* (*φ* = 0.1, 0.6 or 1.0). Results were similar for other values of *τ*, when testing the analysis model with *C* partially observed, and for Scenarios 2-4 (see Supplementary Material, Tables S5-S7 for full results).

**Table 3.** Sensitivity and type 1 error of various tests of linear regression model mis-specification. Results shown for Scenario 1 when testing the analysis model for different strengths of the non-linear association (φ) between X and Y, when the strength of the missingness association (τ) = 1 and Y was partially observed.

| $\phi$ | Test sensitivity (type 1 error) | | | | | |
| --- | --- | --- | --- | --- | --- | --- |
|  | 1. Link | 2. Shapiro-Wilk | 3. Heteroskedasticity | Model for regression of residuals on fitted values |  |  |
|  |  |  |  | 4. FP deg. 2 test | 5. FP deg. 1 test | 6. Grouped regression |
| 0.1 | 0.92 (0.00) | 0.04 (0.05) | 0.05 (0.04) | 0.71 (0.01) | 0.09 (0.04) | 0.33 (0.02) |
| 0.6 | 1.00 (0.01) | 1.00 (0.05) | 0.35 (0.04) | 1.00 (0.03) | 1.00 (0.04) | 1.00 (0.02) |
| 1.0 | 1.00 (0.03) | 1.00 (0.04) | 0.51 (0.05) | 1.00 (0.03) | 1.00 (0.04) | 1.00 (0.02) |

FP, fractional polynomial

**Table 4.** Sensitivity and type 1 error of various tests of logistic regression model mis-specification. Results shown for Scenario 1 when testing the imputation model for C for different strengths of the non-linear association (φ) between X and Y, when the strength of the missingness association (τ) = 1 and C was partially observed.

| $\phi$ | Test sensitivity (type 1 error) | | |
| --- | --- | --- | --- |
|  | 7. Link | 8. Hinkley* | 9. Hosmer-Lemeshow |
| 0.1 | 0.10 (0.03) | 0.03 (0.03) | 0.05 (0.04) |
| 0.6 | 0.81 (0.02) | 0.46 (0.02) | 0.45 (0.04) |
| 1.0 | 0.96 (0.02) | 0.87 (0.01) | 0.73 (0.04) |

\* Hinkley test model did not converge in 3% of simulations

When the non-linear association was weak (*φ* = 0.1), in which case the bias of 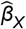 across the analysis approaches was relatively small (see Figure 1, plot I), most tests had low sensitivity to model mis-specification. However, of the linear regression model tests, the link test was highly sensitive even when the model was only slightly mis-specified *i.e*. it highlighted mis-specification when the bias was not practically important. Therefore, in Scenarios 2-4 we used the FP degree-two test rather than the link test for linear regression models (because its sensitivity was more proportional to the bias of 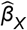). When the non-linear association was stronger (*φ* = 0.6 or 1.0), all linear regression model tests, except the heteroskedasticity test, detected model mis-specification in all simulations (Table 3, sensitivity = 1.00 in each case). Logistic regression tests were less sensitive in these situations, with the link test most sensitive to model mis-specification. Hence, we used the link test for logistic regression models in Scenarios 2-4. Reassuringly, when the analysis or imputation model was correctly specified, type 1 error was ≤ 0.05 for each test.

## 4. Analysis of the motivating example

To illustrate our methods, we used data from the randomised controlled trial described earlier [7, 8]. Here, our focus was on the adjusted relationship between headache score at baseline and headache score at one year follow-up: specifically, whether our estimate of this relationship might be biased by mis-specification of the imputation model.

### 4.1. Methods

We performed a linear regression of headache score at one year on headache score at baseline, adjusting for treatment allocation group (being randomised to receive acupuncture plus standard care, versus receiving standard care alone), and other baseline variables: age, sex, diagnosis (migraine or tension-type headache), and chronicity (number of years of headache disorder). Following the main trial analysis design, we included continuous variables (baseline headache score, age, and chronicity) as linear terms in the analysis model.

Headache score at one year was observed for 301 (75%) of the 401 trial participants. Baseline data were completely observed. We used MI to handle missing values of the headache score at one year. Initially, we used an imputation model for the outcome that was compatible with, but no richer than, the analysis model *i.e*. we used an imputation model identical to the analysis model.

We then assessed possible bias in our estimate of the coefficient of baseline headache score, due to imputation model mis-specification (specifically, incorrectly imposing a linear association for all continuous variables in the imputation model). Since the outcome was continuous, we did this firstly by applying the FP degree-two test (test 4 in Section 3.3) for a linear regression model, and secondly by comparing CRA, MI and PMM estimates. For consistency with the main trial analysis design, we decided not to change the analysis model, but only the imputation model, in light of any detected analysis model mis-specification. To correct any imputation model mis-specification, we identified the best functional form for each continuous covariate in the imputation model. We did this by selecting the best-fitting FP for each continuous variable in turn, when fitting a regression model for headache score at one year that also included all other covariates. We then updated the MI estimates after including any required FP terms in the imputation model. For MI and PMM, we used 25 imputations, reflecting the percentage of participants with a missing outcome [18]. For PMM, we used a donor pool of size five. As per the simulation study, only one variable was partially observed and so no iterations were performed in the imputation procedures. Stata code to perform the real data analysis is included in Supplementary Material, Section S12.

Note that the MI methods considered here are only valid if the outcome is not MNAR. However, a MNAR mechanism is plausible in the context of this trial *e.g*. participants who experienced less severe headaches may have been less motivated to continue to participate in the trial. This issue was explored in the original trial – all participants were contacted at one year and all but 24 provided a global (one-off) estimate of headache severity, which was used in a sensitivity analysis. More recently, Cro *et al*. [19] performed extensive sensitivity analyses using the same data. For simplicity and only for illustration, here we assume the outcome is MAR, given the observed baseline data.

### 4.2. Results

Table 5 shows the estimated mean increase in headache score at one year (conditional on the covariates) per unit increase in baseline headache score, using the different analysis approaches. When analysis and imputation models included only linear terms, CRA and MI estimates were very similar (CRA: 0.71, 95% CI: 0.63-0.79 and MI: 0.70, 95% CI: 0.62-0.79). However, the PMM estimate was slightly larger (0.74, 95% CI: 0.66-0.83). The FP degree-two test suggested that the analysis model was mis-specified (p=0.001). Taken together, these results suggested that CRA and MI estimates were slightly attenuated due to analysis model mis-specification. Specifically, due to the assumption of a linear relationship between continuous covariates and the outcome.

**Table 5.** Mean increase in headache score at one year, per unit increase in baseline headache score, using different analysis approaches

| Analysis approach | Mean increase in headache score at one year, per unit increase in baseline headache score* |  |
| --- | --- | --- |
|  | Estimate | 95% CI |
| CRA | 0.71 | 0.63-0.79 |
| MI including linear terms only | 0.70 | 0.62-0.79 |
| PMM | 0.74 | 0.66-0.83 |
| **After imputation model refinement:** |  |  |
| MI including linear and squared term for baseline headache score | 0.73 | 0.65-0.81 |

\* Adjusted for treatment group, age, sex, diagnosis, and chronicity

Applying FP selection for each continuous covariate in turn suggested a quadratic relationship between baseline headache score and headache score at one year (p=0.002), but no evidence of a non-linear association between age (p=0.530) or chronicity (p=0.409) and headache score at one year. After including a linear and a squared term for baseline headache score in the imputation model (but leaving the analysis model unchanged), the updated MI estimate (0.73, 95% CI: 0.65-0.81) was larger than the CRA and original MI estimates, and closer to the PMM estimate. There was no longer evidence of model mis-specification (p=0.915).

## 5. Discussion

In this paper, we have used a comprehensive simulation study to show that CRA and MI estimates can be biased in situations in which many researchers would expect these approaches to be valid, namely when data are MAR, the analysis and imputation models are compatible, and missingness does not depend on the outcome variable [20].

Our results showed that CRA and MI estimates of the exposure coefficient can be substantially biased if the relationship between the exposure and outcome is mis-specified, or the relationship between the exposure and confounder is mis-specified in the imputation model for the exposure. This is because the (mis-specified) relationship with the exposure in records with missing data differs from the (mis-specified) relationship in records with fully observed data. Hence, the full data value of the exposure coefficient cannot be recovered from the partially observed data (see Supplementary Material, Section 10, for a detailed explanation and illustration). In our simulations, when fitting a linear regression model with no interactions, we found that bias was much smaller if the relationship between the outcome and confounder was mis-specified. However, bias may have been larger in more complex settings *e.g*. if there were interactions, or if fitting a logistic regression model, due to the non-collapsibility property [21].

Further, we found that model mis-specification tests were effective in identifying mis-specified relationships. Such tests can be easily applied to the complete records in any setting in which data are MAR and CRA is valid (*i.e*. when missingness does not depend on the analysis outcome). This can be verified using a “missingness” directed acyclic graph [22] (see Supplementary Material, Section 9, for further details). In settings similar to those in our simulations, we recommend a residual test (based on FP selection) for linear regression model mis-specification, and Pregibon’s link test for logistic regression model mis-specification. Visual inspection of residual plots may additionally be helpful, as this can give hints to the nature of mis-specification. However, model mis-specification may not be visually apparent from the observed data (see, for example, the plots in Supplementary Material, Section 10). Hence, we recommend using statistical tests for model mis-specification, rather than visual inspection alone.

As in previous studies [10, 23], we found that PMM can mitigate for model mis-specification and thus PMM estimates may be substantially different from both CRA and MI estimates in the presence of model mis-specification (note that Supplementary Material Table S1 shows that this is not due to increased variability of PMM point estimates). Therefore, when model mis-specification has been identified, a comparison of CRA, MI, and PMM estimates can be useful for assessing the potential impact of model mis-specification. By contrast, when all models were correctly specified, our results showed CRA and MI estimates were unbiased but PMM estimates had some bias. Therefore, a comparison of estimates is only informative when model mis-specification has been identified by the tests.

In addition to correcting any imputation model mis-specification, a further decision is whether to change the analysis model in light of mis-specification. There may be valid clinical and scientific reasons for retaining linear relationships between all variables in the analysis model (such as pre-specification in a clinical trial setting, or particular interest in the average marginal effect). Any decision to change the analysis model must take into account the study aims, the strength and complexity of the true relationships between variables, the strength of the missingness association, and which variables are partially observed. Our work shows that such decisions are better informed when the imputation model is correctly specified.

Re-analysis of the acupuncture trial showed a non-linear effect of baseline headache score on the primary outcome of headache score at one year follow-up. Consistent with the previous paragraphs, not only was this mis-specification highlighted by the tests, but also by a difference between the PMM and MI analyses. Including the non-linear effect in the imputation model brought the PMM and MI estimates of the coefficient of baseline headache score in the analysis model together. While this does not change the overall interpretation in this example, in other examples this readily avoidable bias could have marked impact.

Note that if all covariates in the analysis model, or all predictors in the imputation model, are binary, then the form of mis-specification we consider here cannot occur *e.g*. if the outcome is continuous and partially observed, and all covariates in the analysis model are binary, bias due to mis-specification of the functional form of the covariates in the analysis model cannot occur (assuming a saturated model *i.e*. that there are no missing higher order interaction terms). However, in this scenario, if the binary exposure is partially observed, then mis-specification of the relationship between the exposure and continuous outcome in the imputation model could result in bias, and should be checked.

A strength of our approach is that we have considered a range of scenarios in which model mis-specification is likely to occur in real data, varying the strengths of both the non-linear and missingness associations. In addition, we have considered a wide range of tests for identifying model mis-specification, in both linear and logistic regression models. In our analyses, we considered scenarios with missing values in a single variable, but our results are generalisable to multivariable missingness (although in this case, assessment of model mis-specification for each partially observed variable in turn may be a more complex process). A limitation of our study (as with any simulation study) is that we have not considered all possible scenarios. In particular, we have only considered mis-specification of the functional form for each variable in the analysis/imputation model. We have not considered other possible model mis-specification, such as not including interactions, mis-specification of the link function in the analysis model, or mis-specification due to using models that are more complex than the true model. However, we would expect our findings to extend to these situations.

We conclude that when using MI methods that assume MAR, compatibility between the analysis and imputation models is necessary, but is not sufficient to avoid bias (or bias amplification). It is important to check (as far as possible) that each imputation model is correctly specified, bearing in mind that an incorrect imputation model can be a consequence of an incorrect analysis model. Our recommended procedure for identifying and correcting any imputation model mis-specification, when performing MI, is outlined in Table 6.

**Table 6.** Procedure for identifying and correcting imputation model mis-specification when using MI.

| Step | Method |
| --- | --- |
| 1 | Test for analysis model mis-specification:<br>i. Fit the analysis model to the complete records (perform a CRA)<br>ii. Perform a model mis-specification test (e.g. regress the residuals on a FP of the fitted values if using a linear regression model, or Pregibon's link test if using a logistic regression model) |
| 2 | If there is evidence of analysis model mis-specification AND any of the partially observed variables are continuous, compare CRA, MI and PMM estimates using the current specification of the analysis model. PMM estimates similar to CRA and MI estimates would suggest analysis model mis-specification has little impact on the estimate of interest.<br>Note that this step should not be applied if model mis-specification is not identified in step 1 (as in this case, differences between the estimates may be due to bias in the PMM estimate). |
| 3 | Identify (as far as possible) the correct specification of the analysis model (e.g. by using FP selection for each continuous covariate in turn). |
| 4 | Decide whether to re-specify the analysis model, or just imputation models, in light of analysis model mis-specification |
| 5 | For each partially observed variable in turn, test for imputation model mis-specification, ensuring that each imputation model is compatible with the corrected analysis model *i.e.* including variables in the same form, and including any interactions implied by the corrected analysis model. |
| 6 | Correct any imputation model mis-specification (e.g. using FP selection for each continuous predictor in turn in the fully conditional specification [25] imputation algorithm) – ensuring that each conditional model remains consistent with the others.<br>Note that steps 5 and 6 do not need to be applied if only the outcome is partially observed and there are no auxiliary variables. |
| 7 | Perform MI using the corrected imputation models (and possibly, a corrected analysis model). |

Procedure assumes that a linear or logistic regression model is fitted, that at least one analysis model covariate/imputation model predictor is continuous, that data are MAR, and that CRA is, in principle, valid.

## Data Availability

Stata code to generate and analyse the data as per the simulation study is included in Supplementary Material, Section S11. Stata code to analyse the real data example is included in Supplementary Material, Section S12. The real data are available at: https://www.ncbi.nlm.nih.gov/pmc/articles/PMC1489946/#S1.

## Acknowledgements

We thank Prof. Andrew Vickers and the acupuncture trial team for the use of their data.

## Notes

### Competing Interest Statement

The authors have declared no competing interest.

### Funding Statement

The results reported herein correspond to specific aims of grant MR/V020641/1 to investigators Kate Tilling and James Carpenter from the UK Medical Research Council. Elinor Curnow, Jon Heron, Rosie Cornish, and Kate Tilling work in the Medical Research Council Integrative Epidemiology Unit at the University of Bristol which is supported by the UK Medical Research Council and the University of Bristol MC_UU_00011/3. James Carpenter is also supported by the UK Medical Research Council (grant no MC_UU_00004/04).

### Author Declarations

Source data were openly available to the public before the initiation of the study. Available at: https://www.ncbi.nlm.nih.gov/pmc/articles/PMC1489946/#S1

