## Supplementary Material for "Multiple imputation of missing data under missing at random: compatible imputation models are not sufficient to avoid bias"

*Table S1. Simulation results for Scenario 1: Y and X are continuous, C is binary, Y depends on C, X, and $X^2$, and X depends on C. Estimates of standardised bias (bias/ $SD(\hat{\beta}_i)$), bias of $\hat{\beta}_i$, $SD(\hat{\beta}_i)$, and model-based standard error (SE) are shown (i = 1 or 2), for complete records analysis (CRA), MI using draws from a linear/logistic imputation model (MI), and MI using type 1 predictive mean matching (PMM). Results are shown for different strengths of the association (τ) between X and missingness of C or Y, and for different strengths of the non-linear association (φ) between X and Y. True values of the exposure and confounder coefficients for the fitted analysis model ($E(Y) = \beta_0 + \beta_1 X + \beta_2 C$) are $\beta_1 = 0.4 + \varphi$, $\beta_2 = 0.8$.*

| **φ** | **Partially observed variable** | **τ** | **Par.** | **Standardised Bias** | | | **Bias of $\hat{\beta}_i$** | | | **SD($\hat{\beta}_i$)** | | | **Model-based SE** | | |
|---|---|---|---|---|---|---|---|---|---|---|---|---|---|---|---|
| | | | | **CRA** | **MI** | **PMM*** | **CRA** | **MI** | **PMM*** | **CRA** | **MI** | **PMM*** | **CRA** | **MI** | **PMM*** |
| 0.1 | C | 0.1 | $\beta_1$ | 0.130 | -0.015 | n/a | 0.005 | -0.001 | n/a | 0.042 | 0.036 | n/a | 0.039 | 0.034 | n/a |
| | | 0.1 | $\beta_2$ | -0.013 | -0.010 | n/a | -0.001 | -0.001 | n/a | 0.085 | 0.084 | n/a | 0.086 | 0.083 | n/a |
| | | 1.0 | $\beta_1$ | 1.534 | 0.089 | n/a | 0.065 | 0.003 | n/a | 0.043 | 0.035 | n/a | 0.041 | 0.034 | n/a |
| | | 1.0 | $\beta_2$ | -0.137 | 0.003 | n/a | -0.012 | 0.000 | n/a | 0.085 | 0.083 | n/a | 0.084 | 0.082 | n/a |
| | | 3.0 | $\beta_1$ | 2.699 | 0.191 | n/a | 0.130 | 0.007 | n/a | 0.048 | 0.035 | n/a | 0.047 | 0.034 | n/a |
| | | 3.0 | $\beta_2$ | -0.146 | 0.054 | n/a | -0.012 | 0.004 | n/a | 0.083 | 0.081 | n/a | 0.083 | 0.081 | n/a |
| | | 5.0 | $\beta_1$ | 3.047 | 0.255 | n/a | 0.154 | 0.009 | n/a | 0.051 | 0.035 | n/a | 0.050 | 0.034 | n/a |
| | | 5.0 | $\beta_2$ | -0.136 | 0.051 | n/a | -0.011 | 0.004 | n/a | 0.085 | 0.083 | n/a | 0.083 | 0.081 | n/a |
| 0.1 | Y | 0.1 | $\beta_1$ | 0.168 | 0.163 | -0.004 | 0.007 | 0.006 | 0.000 | 0.040 | 0.040 | 0.040 | 0.039 | 0.039 | 0.038 |
| | | 0.1 | $\beta_2$ | -0.030 | -0.027 | -0.053 | -0.003 | -0.002 | -0.005 | 0.083 | 0.084 | 0.085 | 0.086 | 0.087 | 0.086 |
| | | 1.0 | $\beta_1$ | 1.540 | 1.535 | 0.281 | 0.064 | 0.064 | 0.013 | 0.042 | 0.042 | 0.046 | 0.041 | 0.041 | 0.038 |
| | | 1.0 | $\beta_2$ | -0.101 | -0.104 | -0.333 | -0.008 | -0.009 | -0.029 | 0.084 | 0.085 | 0.087 | 0.084 | 0.085 | 0.084 |
| | | 3.0 | $\beta_1$ | 2.636 | 2.618 | 0.226 | 0.128 | 0.128 | 0.016 | 0.049 | 0.049 | 0.073 | 0.047 | 0.047 | 0.040 |
| | | 3.0 | $\beta_2$ | -0.147 | -0.148 | -0.684 | -0.012 | -0.012 | -0.060 | 0.082 | 0.082 | 0.088 | 0.083 | 0.083 | 0.083 |
| | | 5.0 | $\beta_1$ | 3.038 | 2.995 | 0.113 | 0.154 | 0.154 | 0.010 | 0.051 | 0.051 | 0.087 | 0.050 | 0.050 | 0.040 |
| | | 5.0 | $\beta_2$ | -0.150 | -0.152 | -0.825 | -0.012 | -0.013 | -0.082 | 0.083 | 0.084 | 0.099 | 0.083 | 0.084 | 0.084 |
| 0.6 | C | 0.1 | $\beta_1$ | 0.473 | -0.016 | n/a | 0.043 | -0.001 | n/a | 0.092 | 0.079 | n/a | 0.054 | 0.047 | n/a |
| | | 0.1 | $\beta_2$ | -0.126 | 0.020 | n/a | -0.014 | 0.002 | n/a | 0.111 | 0.109 | n/a | 0.122 | 0.117 | n/a |

| $\varphi$ | Partially observed variable | $\tau$ | Par. | Standardised Bias | | | Bias of $\widehat{\beta}_j$ | | | SD($\widehat{\beta}_j$) | | | Model-based SE | | |
|---|---|---|---|---|---|---|---|---|---|---|---|---|---|---|---|
| | | | | CRA | MI | PMM* | CRA | MI | PMM* | CRA | MI | PMM* | CRA | MI | PMM* |
| 0.6 | C | 1.0 | $\beta_1$ | 4.552 | 0.037 | n/a | 0.398 | 0.003 | n/a | 0.087 | 0.077 | n/a | 0.054 | 0.045 | n/a |
| | | 1.0 | $\beta_2$ | -0.618 | 0.795 | n/a | -0.067 | 0.099 | n/a | 0.109 | 0.124 | n/a | 0.111 | 0.120 | n/a |
| | | 3.0 | $\beta_1$ | 9.756 | 0.335 | n/a | 0.780 | 0.024 | n/a | 0.080 | 0.072 | n/a | 0.055 | 0.044 | n/a |
| | | 3.0 | $\beta_2$ | -0.791 | 2.008 | n/a | -0.076 | 0.248 | n/a | 0.096 | 0.124 | n/a | 0.097 | 0.121 | n/a |
| | | 5.0 | $\beta_1$ | 11.645 | 0.572 | n/a | 0.934 | 0.042 | n/a | 0.080 | 0.073 | n/a | 0.056 | 0.044 | n/a |
| | | 5.0 | $\beta_2$ | -0.775 | 2.452 | n/a | -0.072 | 0.296 | n/a | 0.093 | 0.121 | n/a | 0.094 | 0.121 | n/a |
| 0.6 | Y | 0.1 | $\beta_1$ | 0.468 | 0.468 | 0.061 | 0.042 | 0.042 | 0.005 | 0.089 | 0.089 | 0.083 | 0.054 | 0.055 | 0.052 |
| | | 0.1 | $\beta_2$ | -0.119 | -0.119 | -0.208 | -0.013 | -0.013 | -0.023 | 0.110 | 0.111 | 0.113 | 0.122 | 0.122 | 0.115 |
| | | 1.0 | $\beta_1$ | 4.498 | 4.489 | 0.920 | 0.394 | 0.394 | 0.073 | 0.088 | 0.088 | 0.079 | 0.054 | 0.054 | 0.049 |
| | | 1.0 | $\beta_2$ | -0.635 | -0.630 | -1.467 | -0.066 | -0.066 | -0.142 | 0.105 | 0.105 | 0.097 | 0.111 | 0.111 | 0.108 |
| | | 3.0 | $\beta_1$ | 9.547 | 9.501 | 1.835 | 0.779 | 0.779 | 0.178 | 0.082 | 0.082 | 0.097 | 0.055 | 0.055 | 0.047 |
| | | 3.0 | $\beta_2$ | -0.827 | -0.816 | -2.260 | -0.079 | -0.079 | -0.199 | 0.096 | 0.097 | 0.088 | 0.097 | 0.097 | 0.101 |
| | | 5.0 | $\beta_1$ | 11.691 | 11.668 | 2.069 | 0.932 | 0.932 | 0.229 | 0.080 | 0.080 | 0.111 | 0.056 | 0.057 | 0.046 |
| | | 5.0 | $\beta_2$ | -0.773 | -0.771 | -2.330 | -0.072 | -0.072 | -0.209 | 0.093 | 0.094 | 0.090 | 0.094 | 0.094 | 0.099 |
| 1.0 | C | 0.1 | $\beta_1$ | 0.531 | -0.028 | n/a | 0.074 | -0.003 | n/a | 0.138 | 0.121 | n/a | 0.075 | 0.065 | n/a |
| | | 0.1 | $\beta_2$ | -0.150 | 0.035 | n/a | -0.022 | 0.005 | n/a | 0.147 | 0.151 | n/a | 0.168 | 0.162 | n/a |
| | | 1.0 | $\beta_1$ | 4.802 | -0.114 | n/a | 0.654 | -0.014 | n/a | 0.136 | 0.119 | n/a | 0.072 | 0.062 | n/a |
| | | 1.0 | $\beta_2$ | -0.772 | 1.172 | n/a | -0.104 | 0.213 | n/a | 0.135 | 0.181 | n/a | 0.148 | 0.173 | n/a |
| | | 3.0 | $\beta_1$ | 10.930 | 0.149 | n/a | 1.299 | 0.017 | n/a | 0.119 | 0.111 | n/a | 0.067 | 0.059 | n/a |
| | | 3.0 | $\beta_2$ | -1.119 | 3.087 | n/a | -0.126 | 0.561 | n/a | 0.113 | 0.182 | n/a | 0.119 | 0.170 | n/a |
| | | 5.0 | $\beta_1$ | 13.439 | 0.484 | n/a | 1.549 | 0.053 | n/a | 0.115 | 0.109 | n/a | 0.066 | 0.060 | n/a |
| | | 5.0 | $\beta_2$ | -1.092 | 4.081 | n/a | -0.115 | 0.679 | n/a | 0.105 | 0.166 | n/a | 0.111 | 0.163 | n/a |
| 1.0 | Y | 0.1 | $\beta_1$ | 0.516 | 0.516 | 0.049 | 0.071 | 0.071 | 0.006 | 0.137 | 0.137 | 0.127 | 0.075 | 0.076 | 0.071 |
| | | 0.1 | $\beta_2$ | -0.149 | -0.145 | -0.214 | -0.021 | -0.021 | -0.032 | 0.144 | 0.145 | 0.150 | 0.168 | 0.169 | 0.154 |
| | | 1.0 | $\beta_1$ | 4.805 | 4.793 | 0.848 | 0.654 | 0.654 | 0.100 | 0.136 | 0.136 | 0.118 | 0.072 | 0.072 | 0.066 |
| | | 1.0 | $\beta_2$ | -0.786 | -0.784 | -1.449 | -0.106 | -0.106 | -0.180 | 0.135 | 0.136 | 0.124 | 0.148 | 0.148 | 0.142 |
| | | 3.0 | $\beta_1$ | 10.850 | 10.786 | 2.177 | 1.297 | 1.297 | 0.274 | 0.120 | 0.120 | 0.126 | 0.067 | 0.068 | 0.060 |

| $\varphi$ | Partially observed variable | $\tau$ | Par. | Standardised Bias | | | Bias of $\hat{\beta}_j$ | | | SD($\hat{\beta}_j$) | | | Model-based SE | | |
|---|---|---|---|---|---|---|---|---|---|---|---|---|---|---|---|
| | | | | **CRA** | **MI** | **PMM*** | **CRA** | **MI** | **PMM*** | **CRA** | **MI** | **PMM*** | **CRA** | **MI** | **PMM*** |
| 1.0 | Y | 3.0 | $\beta_2$ | -1.118 | -1.107 | -2.289 | -0.128 | -0.128 | -0.242 | 0.115 | 0.115 | 0.106 | 0.119 | 0.120 | 0.130 |
| | | 5.0 | $\beta_1$ | 13.248 | 13.205 | 2.613 | 1.550 | 1.550 | 0.368 | 0.117 | 0.117 | 0.141 | 0.066 | 0.067 | 0.057 |
| | | 5.0 | $\beta_2$ | -1.105 | -1.105 | -2.425 | -0.118 | -0.118 | -0.251 | 0.106 | 0.107 | 0.104 | 0.111 | 0.112 | 0.125 |

* PMM only applied in settings in which *Y* was partially observed.
Monte Carlo SE of bias is at most 0.006 for $\beta_1$ and $\beta_2$.

*Table S2. Simulation results for Scenario 2: Y, X, and C are continuous, Y depends on X, C, and $C^2$, and X depends on C.*
*Estimates of standardised bias (bias/ $SD(\hat{\beta}_i)$), bias of $\hat{\beta}_i$, $SD(\hat{\beta}_i)$, and model-based standard error (SE) are shown (i = 1 or 2), for complete records analysis (CRA), MI using draws from a linear imputation model (MI), and MI using type 1 predictive mean matching (PMM).*
*Results are shown for different strengths of the association (τ) between C and missingness of X or Y.*
*True values of the exposure and confounder coefficients for the fitted analysis model ($E(Y) = \beta_0 + \beta_1 X + \beta_2 C$) are $\beta_1 = 0.4$, $\beta_2 = 1.4$.*

| **Partially observed variable** | **τ** | **Par.** | **Standardised Bias** | | | **Bias of $\hat{\beta}_i$** | | | **$SD(\hat{\beta}_i)$** | | | **Model-based SE** | | |
|---|---|---|---|---|---|---|---|---|---|---|---|---|---|---|
| | | | **CRA** | **MI** | **PMM** | **CRA** | **MI** | **PMM** | **CRA** | **MI** | **PMM** | **CRA** | **MI** | **PMM** |
| X | 0.1 | $\beta_1$ | -0.024 | -0.019 | -0.509 | -0.001 | -0.001 | -0.028 | 0.052 | 0.051 | 0.055 | 0.050 | 0.049 | 0.050 |
| | 0.1 | $\beta_2$ | 0.418 | 0.080 | 0.342 | 0.039 | 0.007 | 0.030 | 0.094 | 0.085 | 0.087 | 0.070 | 0.064 | 0.065 |
| | 1.0 | $\beta_1$ | -0.039 | 0.697 | 0.292 | -0.002 | 0.036 | 0.017 | 0.046 | 0.051 | 0.058 | 0.047 | 0.049 | 0.052 |
| | 1.0 | $\beta_2$ | 3.592 | 0.007 | -0.049 | 0.324 | 0.001 | -0.004 | 0.090 | 0.083 | 0.087 | 0.068 | 0.060 | 0.063 |
| | 3.0 | $\beta_1$ | -0.023 | 1.990 | 1.708 | -0.001 | 0.104 | 0.101 | 0.042 | 0.052 | 0.059 | 0.043 | 0.050 | 0.052 |
| | 3.0 | $\beta_2$ | 8.080 | -0.037 | -0.382 | 0.691 | -0.003 | -0.031 | 0.086 | 0.077 | 0.081 | 0.069 | 0.055 | 0.057 |
| | 5.0 | $\beta_1$ | -0.050 | 2.395 | 2.116 | -0.002 | 0.121 | 0.119 | 0.040 | 0.051 | 0.056 | 0.041 | 0.049 | 0.051 |
| | 5.0 | $\beta_2$ | 9.493 | 0.017 | -0.308 | 0.789 | 0.001 | -0.024 | 0.083 | 0.075 | 0.079 | 0.069 | 0.053 | 0.055 |
| Y | 0.1 | $\beta_1$ | 0.012 | 0.009 | -0.133 | 0.001 | 0.000 | -0.007 | 0.051 | 0.051 | 0.052 | 0.050 | 0.050 | 0.047 |
| | 0.1 | $\beta_2$ | 0.392 | 0.392 | 0.095 | 0.037 | 0.037 | 0.009 | 0.093 | 0.094 | 0.090 | 0.070 | 0.071 | 0.067 |
| | 1.0 | $\beta_1$ | -0.017 | -0.020 | -1.120 | -0.001 | -0.001 | -0.051 | 0.047 | 0.047 | 0.046 | 0.047 | 0.047 | 0.045 |
| | 1.0 | $\beta_2$ | 3.571 | 3.567 | 1.079 | 0.324 | 0.324 | 0.090 | 0.091 | 0.091 | 0.083 | 0.068 | 0.069 | 0.064 |
| | 3.0 | $\beta_1$ | -0.045 | -0.045 | -1.825 | -0.002 | -0.002 | -0.081 | 0.043 | 0.043 | 0.044 | 0.043 | 0.043 | 0.044 |
| | 3.0 | $\beta_2$ | 7.937 | 7.882 | 1.694 | 0.691 | 0.692 | 0.165 | 0.087 | 0.088 | 0.097 | 0.069 | 0.070 | 0.064 |
| | 5.0 | $\beta_1$ | -0.032 | -0.033 | -1.900 | -0.001 | -0.001 | -0.084 | 0.041 | 0.041 | 0.044 | 0.041 | 0.042 | 0.044 |
| | 5.0 | $\beta_2$ | 9.104 | 9.042 | 1.618 | 0.787 | 0.786 | 0.178 | 0.086 | 0.087 | 0.110 | 0.069 | 0.069 | 0.063 |

Monte Carlo SE of bias is at most 0.003 for $\beta_1$ and $\beta_2$.

*Table S3. Simulation results for Scenario 3: Y, X, and C are continuous, Y depends on X and C, and X depends on $C^2$.*
*Estimates of standardised bias (bias/ SD($\hat{\beta}_i$)), bias of $\hat{\beta}_i$, SD($\hat{\beta}_i$), and model-based standard error (SE) are shown (i = 1 or 2), for complete records analysis (CRA), MI using draws from a linear imputation model (MI), and MI using type 1 predictive mean matching (PMM).*
*Results are shown for different strengths of the association (τ) between C and missingness of X or Y.*
*True values of the exposure and confounder coefficients for the fitted analysis model ($E(Y) = \beta_0 + \beta_1 X + \beta_2 C$) are $\beta_1 = 0.4$, $\beta_2 = 0.8$.*

| Partially observed variable | τ | Par. | Standardised Bias | | | Bias of $\hat{\beta}_i$ | | | SD($\hat{\beta}_i$) | | | Model-based SE | | |
|---|---|---|---|---|---|---|---|---|---|---|---|---|---|---|
| | | | CRA | MI | PMM | CRA | MI | PMM | CRA | MI | PMM | CRA | MI | PMM |
| X | 0.1 | $\beta_1$ | 0.060 | 0.050 | 0.049 | 0.001 | 0.001 | 0.001 | 0.022 | 0.021 | 0.021 | 0.022 | 0.021 | 0.021 |
| | 0.1 | $\beta_2$ | -0.053 | -0.411 | -0.314 | -0.002 | -0.019 | -0.014 | 0.046 | 0.045 | 0.045 | 0.045 | 0.040 | 0.040 |
| | 1.0 | $\beta_1$ | 0.040 | 0.688 | 0.139 | 0.001 | 0.016 | 0.004 | 0.023 | 0.024 | 0.027 | 0.024 | 0.023 | 0.024 |
| | 1.0 | $\beta_2$ | -0.044 | -3.191 | -1.786 | -0.002 | -0.175 | -0.099 | 0.055 | 0.055 | 0.056 | 0.055 | 0.046 | 0.046 |
| | 3.0 | $\beta_1$ | 0.031 | 1.592 | 0.358 | 0.001 | 0.046 | 0.012 | 0.028 | 0.029 | 0.035 | 0.029 | 0.029 | 0.026 |
| | 3.0 | $\beta_2$ | -0.012 | -6.069 | -2.204 | -0.001 | -0.446 | -0.180 | 0.078 | 0.073 | 0.082 | 0.078 | 0.064 | 0.051 |
| | 5.0 | $\beta_1$ | 0.050 | 1.820 | 0.769 | 0.001 | 0.055 | 0.029 | 0.029 | 0.030 | 0.037 | 0.030 | 0.031 | 0.026 |
| | 5.0 | $\beta_2$ | -0.038 | -6.972 | -2.532 | -0.003 | -0.528 | -0.236 | 0.083 | 0.076 | 0.093 | 0.085 | 0.070 | 0.053 |
| Y | 0.1 | $\beta_1$ | 0.046 | 0.049 | -0.113 | 0.001 | 0.001 | -0.002 | 0.021 | 0.021 | 0.022 | 0.022 | 0.022 | 0.022 |
| | 0.1 | $\beta_2$ | -0.060 | -0.066 | -0.068 | -0.003 | -0.003 | -0.003 | 0.046 | 0.046 | 0.046 | 0.045 | 0.045 | 0.045 |
| | 1.0 | $\beta_1$ | 0.063 | 0.056 | 0.035 | 0.001 | 0.001 | 0.001 | 0.023 | 0.023 | 0.024 | 0.024 | 0.024 | 0.024 |
| | 1.0 | $\beta_2$ | -0.046 | -0.040 | -0.046 | -0.003 | -0.002 | -0.003 | 0.056 | 0.056 | 0.058 | 0.054 | 0.055 | 0.055 |
| | 3.0 | $\beta_1$ | 0.066 | 0.056 | 0.040 | 0.002 | 0.002 | 0.001 | 0.028 | 0.029 | 0.029 | 0.029 | 0.029 | 0.029 |
| | 3.0 | $\beta_2$ | -0.069 | -0.058 | -0.108 | -0.005 | -0.005 | -0.009 | 0.079 | 0.080 | 0.082 | 0.078 | 0.079 | 0.078 |
| | 5.0 | $\beta_1$ | 0.067 | 0.066 | 0.023 | 0.002 | 0.002 | 0.001 | 0.029 | 0.030 | 0.030 | 0.030 | 0.030 | 0.030 |
| | 5.0 | $\beta_2$ | -0.059 | -0.060 | -0.118 | -0.005 | -0.005 | -0.010 | 0.083 | 0.084 | 0.085 | 0.085 | 0.086 | 0.084 |

Monte Carlo SE of bias is at most 0.003 for $\beta_1$ and $\beta_2$.

*Table S4. Simulation results for Scenario 4: Y and C are binary, X is continuous, log-odds of Y depends on $X^2$, and X depends on C. Estimates of standardised bias (bias/ SD($\hat{\beta}_i$)), bias of $\hat{\beta}_i$, SD($\hat{\beta}_i$), and model-based standard error (SE) are shown (i = 1 or 2), for complete records analysis (CRA), and MI using draws from a logistic imputation model (MI).*
*Results are shown for different strengths of the association (τ) between X and missingness of C or Y.*
*True values of the exposure and confounder coefficients for the fitted analysis model (logit{P(Y=1)} = $\beta_0 + \beta_1 X + \beta_2 C$) are $\beta_1 = 0.5$, $\beta_2 = 0.8$.*

| **Partially observed variable** | **τ** | **Par.** | **Standardised Bias** | | **Bias of $\hat{\beta}_i$** | | **SD($\hat{\beta}_i$)** | | **Model-based SE** | |
|---|---|---|---|---|---|---|---|---|---|---|
| | | | **CRA** | **MI** | **CRA** | **MI** | **CRA** | **MI** | **CRA** | **MI** |
| C | 0.1 | $\beta_1$ | 0.478 | 0.228 | 0.044 | 0.018 | 0.092 | 0.077 | 0.091 | 0.078 |
| | 0.1 | $\beta_2$ | 0.140 | 0.159 | 0.025 | 0.028 | 0.178 | 0.178 | 0.187 | 0.188 |
| | 1.0 | $\beta_1$ | 2.571 | 0.278 | 0.280 | 0.022 | 0.109 | 0.078 | 0.106 | 0.078 |
| | 1.0 | $\beta_2$ | -0.030 | 0.129 | -0.005 | 0.024 | 0.185 | 0.184 | 0.188 | 0.189 |
| | 3.0 | $\beta_1$ | 4.637 | 0.320 | 0.598 | 0.024 | 0.129 | 0.076 | 0.137 | 0.078 |
| | 3.0 | $\beta_2$ | -0.024 | 0.198 | -0.004 | 0.037 | 0.185 | 0.186 | 0.189 | 0.188 |
| | 5.0 | $\beta_1$ | 5.099 | 0.334 | 0.726 | 0.025 | 0.142 | 0.076 | 0.154 | 0.078 |
| | 5.0 | $\beta_2$ | -0.024 | 0.210 | -0.004 | 0.039 | 0.186 | 0.185 | 0.191 | 0.190 |
| Y | 0.1 | $\beta_1$ | 0.466 | 0.468 | 0.043 | 0.043 | 0.092 | 0.092 | 0.091 | 0.092 |
| | 0.1 | $\beta_2$ | 0.125 | 0.124 | 0.023 | 0.023 | 0.184 | 0.185 | 0.187 | 0.188 |
| | 1.0 | $\beta_1$ | 2.720 | 2.715 | 0.283 | 0.283 | 0.104 | 0.104 | 0.107 | 0.108 |
| | 1.0 | $\beta_2$ | 0.000 | -0.003 | 0.000 | -0.001 | 0.185 | 0.185 | 0.189 | 0.190 |
| | 3.0 | $\beta_1$ | 4.583 | 4.550 | 0.599 | 0.597 | 0.131 | 0.131 | 0.136 | 0.138 |
| | 3.0 | $\beta_2$ | -0.021 | -0.022 | -0.004 | -0.004 | 0.187 | 0.188 | 0.189 | 0.189 |
| | 5.0 | $\beta_1$ | 5.041 | 5.004 | 0.728 | 0.726 | 0.144 | 0.145 | 0.154 | 0.155 |
| | 5.0 | $\beta_2$ | -0.011 | -0.006 | -0.002 | -0.001 | 0.186 | 0.186 | 0.191 | 0.192 |

Monte Carlo SE of bias is at most 0.006 for $\beta_1$ and $\beta_2$.

*Table S5. Sensitivity and type 1 error of various tests of analysis model mis-specification when fitting a linear regression model. Results are shown for scenario 1 when either C or Y were partially observed, for different strengths of the association (τ) between X and missingness of C or Y, and for different strengths of the non-linear association (φ) between X and Y.*

| **φ** | **Partially observed variable** | **τ** | **Test sensitivity (type 1 error)** | | | | | | | | | | | |
|---|---|---|---|---|---|---|---|---|---|---|---|---|---|---|
| | | | | | | | | | **Model for regression of residuals on fitted values** | | | | | |
| | | | **1. Link** | | **2. Shapiro-Wilk** | | **3. Hetero-skedasticity** | | **4. FP deg. 2 test** | | **5. FP deg. 1 test** | | **6. Grouped regression** | |
| 0.1 | *C* | 0.1 | 0.97 | (0.00) | 0.05 | (0.05) | 0.06 | (0.04) | 0.85 | (0.02) | 0.10 | (0.04) | 0.46 | (0.01) |
| | | 1 | 0.93 | (0.00) | 0.05 | (0.05) | 0.05 | (0.05) | 0.71 | (0.02) | 0.05 | (0.05) | 0.30 | (0.01) |
| | | 3 | 0.64 | (0.00) | 0.05 | (0.04) | 0.06 | (0.05) | 0.31 | (0.01) | 0.04 | (0.04) | 0.11 | (0.01) |
| | | 5 | 0.48 | (0.00) | 0.05 | (0.04) | 0.06 | (0.05) | 0.18 | (0.02) | 0.03 | (0.04) | 0.08 | (0.01) |
| | *Y* | 0.1 | 0.97 | (0.00) | 0.06 | (0.06) | 0.08 | (0.05) | 0.86 | (0.02) | 0.10 | (0.04) | 0.45 | (0.01) |
| | | 1 | 0.92 | (0.00) | 0.04 | (0.05) | 0.05 | (0.04) | 0.71 | (0.01) | 0.09 | (0.04) | 0.33 | (0.02) |
| | | 3 | 0.66 | (0.00) | 0.05 | (0.04) | 0.05 | (0.06) | 0.33 | (0.01) | 0.04 | (0.02) | 0.13 | (0.01) |
| | | 5 | 0.49 | (0.00) | 0.04 | (0.04) | 0.05 | (0.05) | 0.16 | (0.01) | 0.03 | (0.03) | 0.07 | (0.02) |
| 0.6 | *C* | 0.1 | 1.00 | (0.02) | 1.00 | (0.05) | 0.36 | (0.05) | 1.00 | (0.04) | 1.00 | (0.05) | 1.00 | (0.02) |
| | | 1 | 1.00 | (0.01) | 1.00 | (0.04) | 0.34 | (0.05) | 1.00 | (0.04) | 1.00 | (0.04) | 1.00 | (0.01) |
| | | 3 | 1.00 | (0.00) | 0.92 | (0.05) | 0.62 | (0.04) | 1.00 | (0.03) | 1.00 | (0.03) | 1.00 | (0.01) |
| | | 5 | 1.00 | (0.00) | 0.70 | (0.04) | 0.78 | (0.04) | 1.00 | (0.02) | 1.00 | (0.04) | 1.00 | (0.01) |
| | *Y* | 0.1 | 1.00 | (0.02) | 1.00 | (0.05) | 0.35 | (0.04) | 1.00 | (0.03) | 1.00 | (0.04) | 1.00 | (0.02) |
| | | 1 | 1.00 | (0.01) | 1.00 | (0.05) | 0.35 | (0.04) | 1.00 | (0.03) | 1.00 | (0.04) | 1.00 | (0.02) |
| | | 3 | 1.00 | (0.00) | 0.92 | (0.05) | 0.60 | (0.04) | 1.00 | (0.03) | 1.00 | (0.03) | 1.00 | (0.01) |
| | | 5 | 1.00 | (0.00) | 0.72 | (0.05) | 0.78 | (0.04) | 1.00 | (0.01) | 1.00 | (0.03) | 1.00 | (0.01) |
| 1.0 | *C* | 0.1 | 1.00 | (0.05) | 1.00 | (0.04) | 0.51 | (0.05) | 1.00 | (0.05) | 1.00 | (0.04) | 1.00 | (0.03) |
| | | 1 | 1.00 | (0.03) | 1.00 | (0.05) | 0.54 | (0.04) | 1.00 | (0.04) | 1.00 | (0.03) | 1.00 | (0.01) |
| | | 3 | 1.00 | (0.00) | 1.00 | (0.04) | 0.85 | (0.04) | 1.00 | (0.03) | 1.00 | (0.03) | 1.00 | (0.01) |
| | | 5 | 1.00 | (0.00) | 1.00 | (0.04) | 0.98 | (0.05) | 1.00 | (0.02) | 1.00 | (0.04) | 1.00 | (0.01) |
| | *Y* | 0.1 | 1.00 | (0.03) | 1.00 | (0.04) | 0.53 | (0.05) | 1.00 | (0.04) | 1.00 | (0.04) | 1.00 | (0.01) |
| | | 1 | 1.00 | (0.03) | 1.00 | (0.04) | 0.51 | (0.05) | 1.00 | (0.03) | 1.00 | (0.04) | 1.00 | (0.02) |
| | | 3 | 1.00 | (0.01) | 1.00 | (0.05) | 0.85 | (0.04) | 1.00 | (0.04) | 1.00 | (0.04) | 1.00 | (0.01) |
| | | 5 | 1.00 | (0.00) | 1.00 | (0.04) | 0.97 | (0.05) | 1.00 | (0.03) | 1.00 | (0.04) | 1.00 | (0.01) |

*Table S6. Sensitivity and type 1 error of various tests of imputation model mis-specification when fitting a logistic regression model, with C partially observed. Results are shown for scenario 1, for different strengths of the association (τ) between X and missingness of C, and for different strengths of the non-linear association (φ) between X and Y.*

| **φ** | **τ** | **Test sensitivity (type 1 error)** | | | | | |
|---|---|---|---|---|---|---|---|
| | | **7. Link** | | **8. Hinkley*** | | **9. Hosmer-Lemeshow** | |
| 0.1 | 0.1 | 0.10 | (0.03) | 0.03 | (0.03) | 0.07 | (0.03) |
| | 1 | 0.10 | (0.03) | 0.03 | (0.03) | 0.05 | (0.04) |
| | 3 | 0.06 | (0.04) | 0.03 | (0.03) | 0.04 | (0.03) |
| | 5 | 0.07 | (0.03) | 0.03 | (0.03) | 0.05 | (0.04) |
| 0.6 | 0.1 | 0.86 | (0.02) | 0.58 | (0.02) | 0.48 | (0.04) |
| | 1 | 0.81 | (0.02) | 0.46 | (0.02) | 0.45 | (0.04) |
| | 3 | 0.62 | (0.03) | 0.24 | (0.02) | 0.26 | (0.04) |
| | 5 | 0.51 | (0.03) | 0.17 | (0.02) | 0.19 | (0.06) |
| 1.0 | 0.1 | 0.98 | (0.02) | 0.91 | (0.01) | 0.78 | (0.03) |
| | 1 | 0.96 | (0.02) | 0.87 | (0.01) | 0.73 | (0.04) |
| | 3 | 0.90 | (0.03) | 0.68 | (0.02) | 0.58 | (0.04) |
| | 5 | 0.85 | (0.03) | 0.53 | (0.02) | 0.46 | (0.04) |

* Hinkley test model did not converge in 3% of simulations

*Table S7. Sensitivity and type 1 error of tests of analysis model mis-specification when Y (continuous in scenarios 2 and 3, and binary in scenario 4) is partially observed, and of imputation model mis-specification when continuous X or binary C are partially observed (test for continuous variables: fitting a degree two fractional polynomial in the regression of the residuals on the fitted values; test for binary variables: Pregibon's link test). Results are shown for scenarios 2 – 4, for different strengths of the missingness association (τ).*
*Note that, because scenario 1 results for analysis model tests were so similar when either Y or C were partially observed, in all other scenarios we only tested the analysis model when Y was partially observed.*

| **Scenario** | **Partially observed variable** | ***τ*** | **Test sensitivity (type 1 error)** | |
|---|---|---|---|---|
| 2 | Y (continuous) | 0.1 | 1.00 | (0.01) |
| | | 1 | 1.00 | (0.01) |
| | | 3 | 1.00 | (0.01) |
| | | 5 | 1.00 | (0.01) |
| | X (continuous) | 0.1 | 1.00 | (0.01) |
| | | 1 | 1.00 | (0.01) |
| | | 3 | 1.00 | (0.01) |
| | | 5 | 1.00 | (0.01) |
| 3 | Y* (continuous) | 0.1 | 0.01 | (0.01) |
| | | 1 | 0.02 | (0.02) |
| | | 3 | 0.02 | (0.02) |
| | | 5 | 0.02 | (0.02) |
| | X (continuous) | 0.1 | 1.00 | (0.01) |
| | | 1 | 1.00 | (0.01) |
| | | 3 | 1.00 | (0.01) |
| | | 5 | 1.00 | (0.01) |
| 4 | Y (binary) | 0.1 | 1.00 | (0.05) |
| | | 1 | 1.00 | (0.04) |
| | | 3 | 0.80 | (0.02) |
| | | 5 | 0.54 | (0.02) |
| | C (binary) | 0.1 | 0.07 | (0.00) |
| | | 1 | 0.04 | (0.00) |
| | | 3 | 0.03 | (0.00) |
| | | 5 | 0.03 | (0.00) |

* In scenario 3, the chosen analysis model correctly specifies the relationship between *Y, X* and *C*. Hence, test sensitivity equals type 1 error in this case.

*Section S8. Description of model mis-specification tests*

**Linear regression model mis-specification tests**

We applied the following tests, designed to assess the validity of the linear regression model:

1. Pregibon [1] (link) test that the correct link function has been used. This was originally designed as a test of the specification of the outcome. However, a large test statistic can also indicate mis-specification of the covariate model.
2. Shapiro-Wilk [2] test of normality of residuals.
3. Breusch–Pagan/Cook–Weisberg [3, 4] (heteroskedasticity) test of constant variance of residuals.
4. Fractional polynomial (FP) degree-two test of no association between residuals and the best-fitting FP of the fitted values. The best-fitting FP is identified using FP selection [5], choosing from a set of polynomial functions based on the model deviance. In degree two FP selection, a pair of polynomials is fitted (choosing the best-fitting pair from the set of all possible pairwise combinations). We chose the fractional powers from the set {–2, –1, –0.5, 0, 0.5, 1, 2, 3, where a power of zero is the log function}.
5. FP degree-one test of no association between residuals and fitted values. This is a variant of test 4, fitting a single polynomial instead of a pair of polynomials.
6. The grouped residuals test fits a regression of the model residuals on the grouped fitted values (grouped according to the quintiles of the distribution of the fitted values).

**Logistic regression model diagnostic tests**

7. Pregibon [1] (link) test that the correct link function has been used. This test can be used for logistic as well as linear regression models.
8. A test based on Hinkley's [6] method. This involves comparing the original fitted model to a model including a FP of the logit of the predicted probabilities from the original fitted model (*i.e.* a FP of the linear predictor).
9. Hosmer-Lemeshow [7] test. This test compares observed and expected event rates in grouped data, grouped according to quantiles of the predicted probabilities. We used ten groups.

*Section S9. Validity of model checking using complete records*

In our simulation scenarios, if the analysis and imputation models were correctly specified, CRA estimates would be unbiased (regardless of the sample size) because missingness does not depend on the outcome *Y*, given the observed data. This can be verified by inspecting the "missingness" directed acyclic graph (DAG) [8] for each scenario (Figure S1).

*Figure S1. For scenarios 1 to 4 in the simulation study, directed acyclic graphs depicting the relationship between Y, X, C, and missingness indicator $R_\Delta$ ($\Delta$ = Y or C in Scenarios 1 and 4; $\Delta$ = Y or X in Scenarios 2 and 3) in CRA.*

*Lines indicate related variables, with arrows indicating the direction of the relationship; absent lines represent conditional independencies. Boxes indicate variables conditioned on in CRA.*

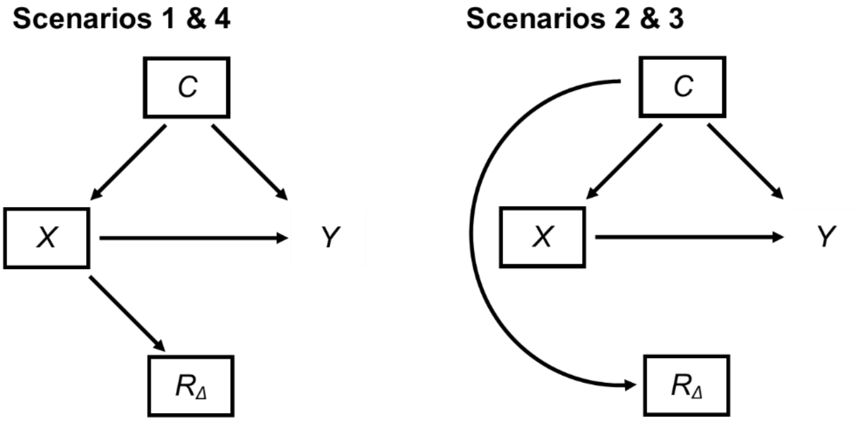


In each DAG in Figure S1, solid lines indicate the relationships between *Y*, *X*, *C*, and missingness indicator $R_\Delta$ ($\Delta = Y$ or *C* in Scenarios 1 and 4; $\Delta = Y$ or *X* in Scenarios 2 and 3), with arrows indicating the direction of the relationship. Boxes indicate the variables conditioned on in CRA. Boxes around *X* and *C* indicate that these variables are regressors in the analysis model. The box around $R_\Delta$ indicates that we only include complete records in CRA.

In all scenarios, missingness does not depend on *Y* (*i.e.* there is no open path between *Y* and $R_\Delta$), given *X* and *C*. Hence, CRA is valid (though not necessarily efficient). In particular, this means that it is valid to use the complete records to check for analysis model mis-specification (assuming positivity). If the model is incorrect, the complete records provide evidence to reject it, provided the number of complete records is large enough (*i.e.* given sufficient power), relative to the severity of mis-specification.

Similarly, if the analysis and imputation models were correctly specified, MI estimates would be unbiased because data are MAR. When *Y* is partially observed, the analysis model and the imputation model are the same (in the absence of auxiliary data), and hence it is not necessary to perform additional tests for mis-specification of the imputation model for *Y*. When *C* or *X* are partially observed (in Scenarios 1 & 4, and 2 & 3, respectively), both *C* and *X* are independent of $R_{\Delta}$, given the variables in their respective imputation models. Again, this could be verified by inspecting missingness DAGs for the imputation models for *C* or *X* (note that these would contain the same variables and relationships as in Figure S1, but the variables conditioned on would differ: we would condition on *X* and *Y* in the imputation model for *C*, and on *C* and *Y* in the imputation model for *X*. To avoid repetition, these DAGs are not shown). Thus, it is also valid to perform model checks for the imputation model for *C* or *X* using the complete records.

Note that in analysis of real data, there may be additional, unmeasured variables related to *Y*, *X*, *C*, and their missingness (in our simulation studies, we have assumed that all required variables are at least partially observed). There may also be several partially observed variables and/or missingness may depend on more than one variable. In this case, checks for analysis and imputation model mis-specification will be more complex, but we can still use missingness DAGs to establish whether it is valid to check for model mis-specification using the complete records. If the missingness DAG indicates that this is not valid (*e.g.* because *Y* is MNAR), we cannot use complete records to identify model mis-specification (and in any case, changing the functional form of variables in the analysis and/or imputation models will not mitigate for data MNAR).

*Section S10. Explanation of bias in CRA and MI estimates in simulation study scenarios*

In Figures S2-S4, we use plots of the simulated data to provide an intuitive explanation for why CRA and/or MI estimates of $\beta_X$ are biased in some of our simulation scenarios. Each plot uses data from 200 randomly selected simulated records. In each plot, whether a record has missing data or not is indicated by hollow or filled circles, respectively. Figure S2 shows plots of *Y* against *X* for Scenario 1, for different strengths of the non-linear association ($\varphi$) between *Y* and *X*, and for different strengths of the association ($\tau$) between missingness (of *Y* or *C*) and *X*. The regression line for the regression of *Y* on *X*, assuming a linear relationship, is also shown for the full data analysis and for CRA.

*Figure S2. Plots of Y against X for Scenario 1, for different strengths of the non-linear association (φ) between Y and X, and for different strengths of the association (τ) between missingness and X. Regression lines assume a linear relationship between Y and X.*

*Each plot uses data from 200 randomly selected simulated records.*

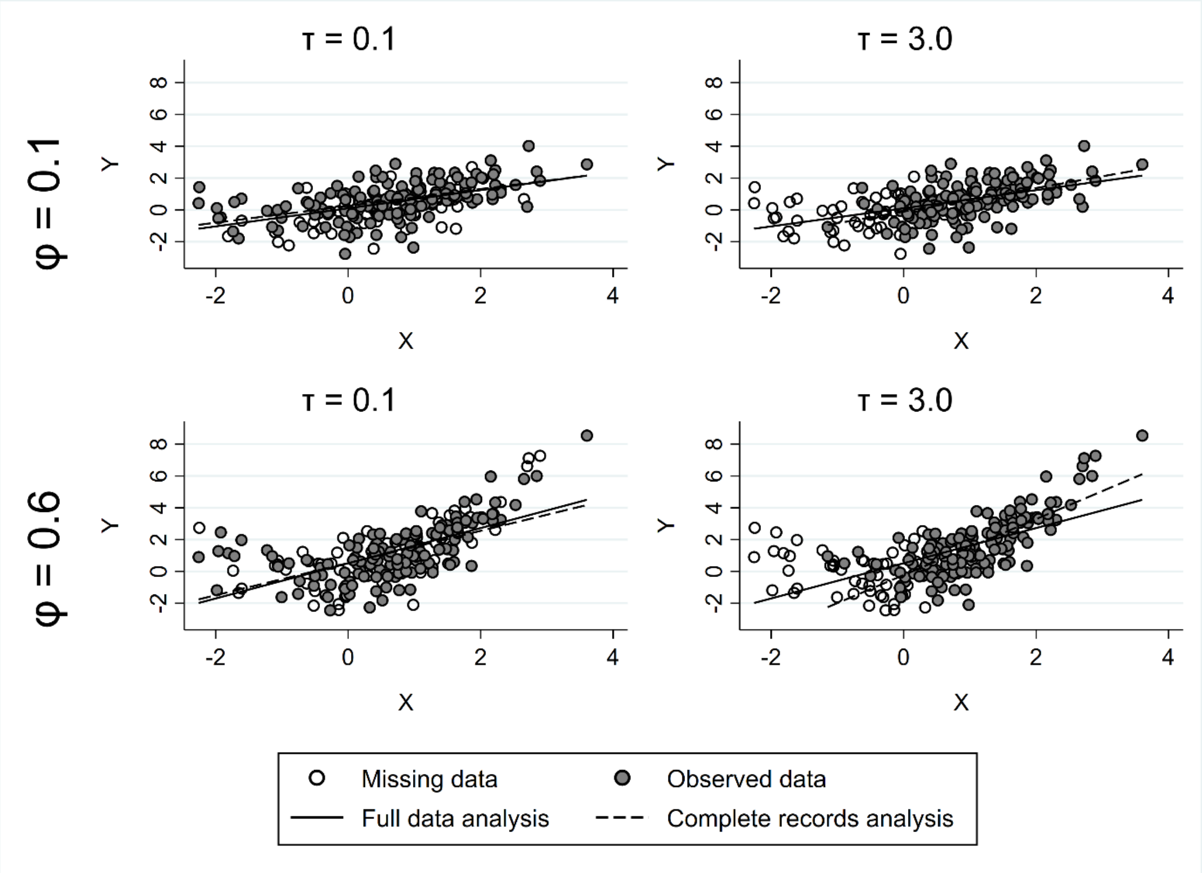


From the left-hand plots in Figure S2, we can see that when data are weakly MAR (when $\tau$ = 0.1), the CRA exposure coefficient (*i.e.* the slope) is similar to the full data coefficient, even when the non-linear association is fairly strong (when $\varphi$ = 0.6). However, when data are strongly MAR (when $\tau$ = 3.0) and the non-linear association is fairly strong (when $\varphi$ = 0.6), records with missing data have a different linear *X*-*Y* relationship from records with fully observed data. Hence, the true (full data) value of the coefficient cannot be recovered from the observed data, neither in CRA, nor in MI. Note this is not the case when a quadratic (the correctly specified relationship) is fitted (Figure S3). The same argument can be used to explain the bias in both CRA and MI estimates in Scenario 4, in which there is also a non-linear relationship between *Y* and X, but *Y* is binary rather than continuous.

*Figure S3. Plot of Y against X for Scenario 1, when the non-linear association (φ) between Y and X = 0.6, and the strength of the association (τ) between missingness and X = 3.0. The regression line assumes a quadratic relationship between X and Y.*

*The plot uses data from 200 randomly selected simulated records.*

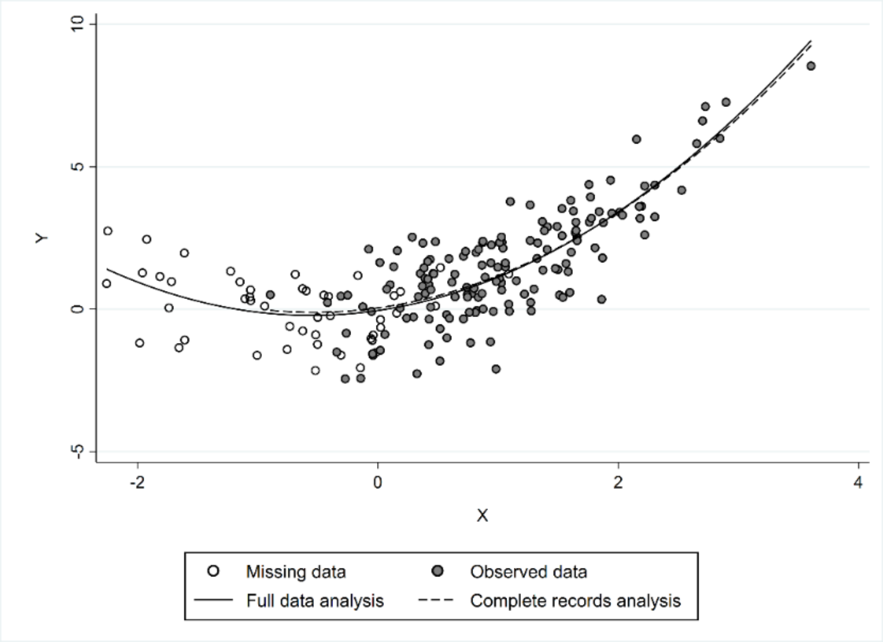


Conversely, Figure S4 (overleaf) shows plots of *Y* against *X* for Scenarios 2 and 3, when data are strongly MAR (when $\tau$ = 3.0). We can see that the CRA regression coefficient (*i.e.* the slope) is similar to the full data coefficient because the relationship between *X* and *Y* is correctly specified in these scenarios. Hence, the true (full data) value of the exposure coefficient can be recovered from the observed data in both CRA, and MI when *Y* is partially observed (although not in MI when *X* is partially observed, because the relationship between *X* and *C* is mis-specified in the imputation model for *X*).

*Figure S4. Plots of Y against X for Scenarios 2 and 3, when the strength of the association (τ) between missingness and C = 3.0. Regression lines assume a linear relationship between X and Y.*

*Each plot uses data from 200 randomly selected simulated records*

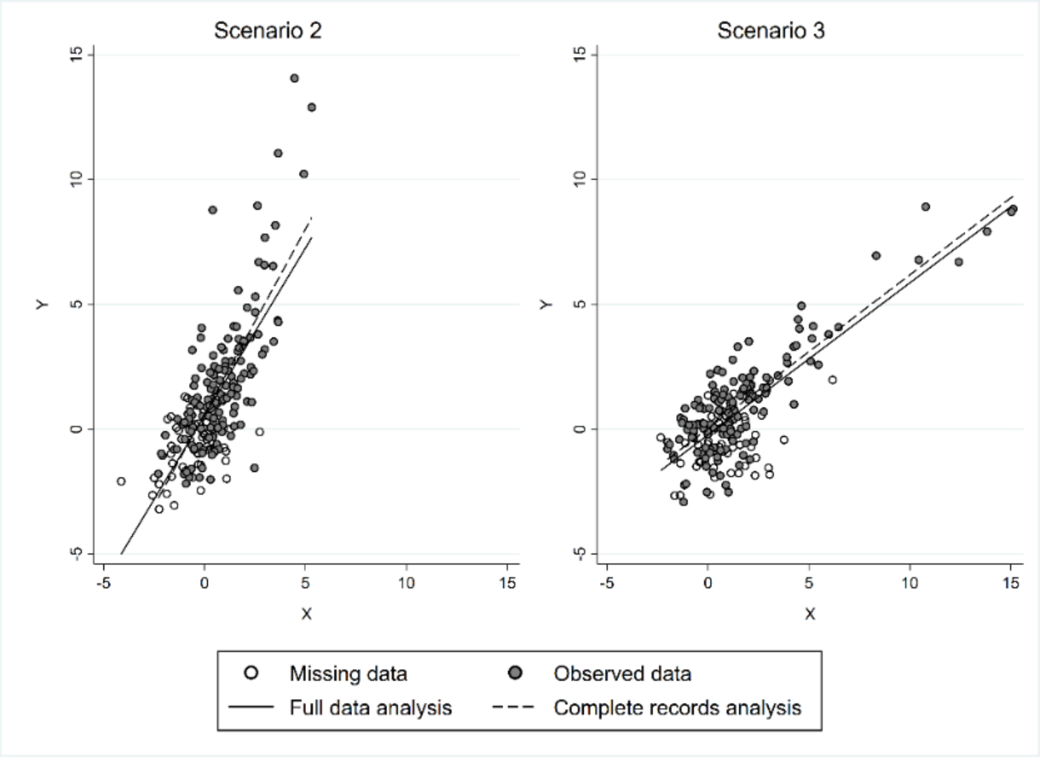

*Section S11. Stata code for the simulation study*

```
****Data generation for sim study********

*Define postfile to store results
tempname simloop
postfile `simloop' float(sampno c3bin c3cts x1_b x1_c x1_d y1_b1_p1
y1_b1_p6 y1_b1_1 y1_c1 y1_d1 y1_b1_bin) using "sim1_DGM1000.dta",
replace

*Also store rngstate
tempname simseed
postfile `simseed' str2000(s1 s2 s3) using
"sim1_DGM1000_seedfile.dta", replace

*Create a temporary file for storing results for each iteration
tempfile tmpDGM

forvalues i=1/1000 {
      *record seed at start of each iteration
      post `simseed' (substr(c(rngstate),1,2000))
(substr(c(rngstate),2001,2000)) (substr(c(rngstate),4001,.))

      clear
      quietly set obs 1000

      *Sample no
      gen sampno=`i'

      gen c3bin=rbinomial(1,0.5)
      gen c3cts=rnormal(0.5,1)

      *Specify DGM for X: either (i) X1 = C3 + ε (C3 either binary
or cts) or X1 = C3^2 + ε, where ε ~ N(0,1)
      gen x1_b=rnormal(c3bin,1)
      gen x1_c=rnormal(c3cts,1)
      gen x1_d=rnormal(c3cts^2,1)

      ****** Scenario 1 - Y ~ N(-0.4 + 0.4 X + 0.8 C3 + phi X^2,1)
      gen y1_b1_p1=rnormal(-0.4 + 0.4*x1_b + 0.8*c3bin +
0.1*x1_b^2,1)
      gen y1_b1_p6=rnormal(-0.4 + 0.4*x1_b + 0.8*c3bin +
0.6*x1_b^2,1)
      gen y1_b1_1=rnormal(-0.4 + 0.4*x1_b + 0.8*c3bin +
1.0*x1_b^2,1)

      ****** Scenario 2 - Y ~ N(-0.4 + 0.4 X + 0.8 C3 + 0.6 C3^2,1)
      gen y1_c1=rnormal(-0.4 + 0.4*x1_c + 0.8*c3cts + 0.6*c3cts^2,1)

      ****** Scenario 3 - Y ~ N(-0.4 + 0.4 X + 0.8 C3,1) ******
      gen y1_d1=rnormal(-0.4 + 0.4*x1_d + 0.8*c3cts,1)

      ****** Scenario 4 - Y ~logit-1 (-0.4 + 0.4 X + 0.6 X2 + 0.8 C)
      gen y1_b1_bin=rbinomial(1,invlogit(-0.4 + 0.4*x1_b + 0.8*c3bin
+ 0.5*x1_b^2))
```

```
      quietly save `tmpDGM', replace
      use sim1_DGM1000, clear
      append using `tmpDGM'
      quietly save sim1_DGM1000, replace
}
postclose `simseed'
postclose `simloop'
```

********Run simulation study********

```
*To avoid repetition, code is supplied for Scenario 1 only
*Code is easily adaptable for Scenarios 2-4

*Define postfile to store results
tempname simloop
postfile `simloop' int(i) float(tau phi) str7(correct dgm_y dgm_x
missvar missmethod) ///
float(const se_const beta_x se_x beta_c3 se_c3 beta_xsq se_xsq link
swilk het FP_d2 FP_d1 quint link_lgt hinkley HL) using
"sim1_yctsB1.dta", replace

*Also store rngstate
tempname simseed
postfile `simseed' str2000(s1 s2 s3) using
"sim1_yctsB1_seedfile.dta", replace

*Create a temporary file for storing imputed data
tempfile tmpfull

forvalues i=1/1000 {
      di "`i'"
      *record seed at start of each iteration
      post `simseed' (substr(c(rngstate),1,2000))
(substr(c(rngstate),2001,2000)) (substr(c(rngstate),4001,.))

      foreach phi of numlist 1.0 0.6 0.1  {

      foreach tau of numlist 5 3 1 0.1 {

*********SCENARIO 1. *****************
* First using correctly specified models to calculate type 1 error
* Code shown only for settings in which C is partially observed
* Code is very similar when Y is partially observed

      quietly use sim1_DGM1000 if sampno==`i', clear
      quietly gen y=y1_b1_p1 if `phi’ < 0.2
      quietly replace y=y1_b1_p6 if `phi’ > 0.2 & `phi’ < 0.7
      quietly replace y=y1_b1_1 if `phi’ > 0.7
      gen x=x1_b
      gen xsq=x1_b^2
      if `tau' < 0.2 local alpha = 8
         else if `tau' > 0.2 & `tau' < 2 local alpha = 0.6
         else if `tau' > 2 & `tau' < 4 local alpha = 0.2
         else local alpha = 0.1
```

```
     quietly gen r_c3=rbinomial(1,invlogit(`tau'*(`alpha' + x)))
* Allowable range for p is 1e-8 to 1-1e-8, so extreme values of x
give missing values for r_c3
     quietly replace r_c3=0 if invlogit(`tau'*(`alpha' + x)) <
0.00000001
     gen c3miss=c3bin
     quietly replace c3miss=. if r_c3==0

     *Diagnostic tests
     *Tests for analysis model mis-specification (linear
regression)
     quietly regress y x c3miss xsq
     *store values for -post-
     est store vals

     *Link test
     quietly linktest
     local link=(2 * ttail(r(df), abs(r(t))))
     * Shapiro-Wilks
     quietly swilk r
     local swilk=r(p)
     *Test for heteroskedasticity
     quietly estat hettest
     local het=r(p)
     * FP tests
     *first using default of dim(2)
     quietly predict r,resid
     quietly predict fit,xb
     quietly fp <fit>, scale: regress r <fit>
     local fp_d2=e(fp_compare)[1,4]
     *then using dim(1)
     drop fit_1 fit_2
     quietly fp <fit>, dim(1) scale: regress r <fit>
     local fp_d1=e(fp_compare)[1,4]
     *Grouped test
     xtile fit_q = fit, nq(5)
     quietly anova r fit_q
     local quint=Ftail(e(df_m),e(df_r),e(F))

     *Tests for imputation model mis-specification (logistic
regression)
     quietly logistic c3miss y x xsq
     *Link test
     quietly linktest
     local link_lgt=(2 *  (1-normal(abs(r(t)))))
     *Hinkley test – using capture as does not always converge
     drop fit fit_1 fit_q
     quietly predict fit,xb
     capture {
     quietly fp <fit>, scale: logit c3miss <fit> y x xsq
     *Compare to model without fit
     local hinkley=e(fp_compare)[1,4]
     }
     *If error, run rest of loop
     if _rc!=0 {
         local hinkley=.
```

```
        }
      *Hosmer-Lemeshow
      quietly estat gof, group(10)
      local HL=r(p)

      *Restore regression estimates (for info) and post all ests
      quietly est restore vals
      post `simloop' (`i') (`tau') (`phi') ("Yes") ("B1") ("X1")
("C") ("CRA") ///
      (_b[_cons]) (_se[_cons]) (_b[x]) (_se[x]) (_b[c3miss])
(_se[c3miss]) ///
      (_b[xsq]) (_se[xsq]) (`link') (`swilk') (`het') (`fp_d2')
(`fp_d1') (`quint') (`link_lgt') (`hinkley') (`HL')


      *****USING INCORRECT MODELS****************
      *1. Missingness in outcome Y
      drop _all
      quietly use sim1_DGM1000 if sampno==`i', clear
      gen x=x1_b
      if `tau' < 0.2 local alpha = 8
         else if `tau' > 0.2 & `tau’ < 2 local alpha = 0.6
         else if `tau' > 2 & `tau’ < 4 local alpha = 0.2
         else local alpha = 0.1
      quietly gen r_y=rbinomial(1,invlogit(`tau'*(`alpha' + x)))
* Allowable range for p is 1e-8 to 1-1e-8, so extreme values of x
give missing values for r_y
      quietly replace r_y=0 if invlogit(`tau'*(`alpha' + x)) <
0.00000001
      quietly gen ymiss=y1_b1_p1 if `phi' < 0.2
      quietly replace ymiss=y1_b1_p6 if `phi' > 0.2 & `phi' < 0.7
      quietly replace ymiss=y1_b1_1 if `phi' > 0.7

      quietly replace ymiss=. if r_y==0
      *save for use with PMM later
      quietly save `tmpfull', replace

      *CRA
      quietly regress ymiss x c3bin
      *store values for -post-
      est store vals

      *Tests for analysis model mis-specification (linear
regression) (using CRA model)

      *Link test
      quietly linktest
      local link=(2 * ttail(r(df), abs(r(t))))
      * Shapiro-Wilks
      quietly swilk r
      local swilk=r(p)
      *Test for heteroskedasticity
      quietly estat hettest
      local het=r(p)
      * FP tests
      quietly predict r,resid
```

```
      quietly predict fit,xb
      *first using default of dim(2)
      quietly fp <fit>, scale: regress r <fit>
      local fp_d2=e(fp_compare)[1,4]
      *then using dim(1)
      drop fit_1 fit_2
      quietly fp <fit>, dim(1) scale: regress r <fit>
      local fp_d1=e(fp_compare)[1,4]
      *Grouped test
      xtile fit_q = fit, nq(5)
      quietly anova r fit_q
      local quint=Ftail(e(df_m),e(df_r),e(F))

      *Restore regression estimates and post all ests
      quietly est restore vals
      post `simloop' (`i') (`tau') (`phi') ("No") ("B1") ("X1")
("Y") ("CRA") ///
      (_b[_cons]) (_se[_cons]) (_b[x]) (_se[x]) (_b[c3bin])
(_se[c3bin]) ///
      (0) (0) (`link') (`swilk') (`het') (`fp_d2') (`fp_d1')
(`quint') (0) (0) (0)

      *** MI linear imputation model ***
      quietly mi set flong
      quietly mi register imputed ymiss
      quietly mi register regular x c3bin
      quietly mi impute chained (regress) ymiss = x c3bin, add(30)
      quietly mi estimate: regress ymiss x c3bin
      post `simloop' (`i') (`tau') (`phi') ("No") ("B1") ("X1")
("Y") ("MI") ///
      (e(b_mi)[1,3]) (sqrt(e(V_mi)[3,3])) (e(b_mi)[1,1])
(sqrt(e(V_mi)[1,1])) ///
      (e(b_mi)[1,2]) (sqrt(e(V_mi)[2,2])) ///
      (0) (0) (0) (0) (0) (0) (0) (0) (0) (0) (0)

      *** PMM ****
      *Note, type 1 PMM can only be run in ice not mi impute
      *restore simulated data i.e. before imputation
      use `tmpfull', clear
      quietly ice ymiss x c3bin, saving(`tmpfull', replace) m(30)
match matchpool(5) uvisopts(matchtype(1))
      use `tmpfull', clear
      quietly mi import ice, automatic
      quietly mi estimate: regress ymiss x c3bin
      post `simloop' (`i') (`tau') (`phi') ("No") ("B1") ("X1")
("Y") ("PMM") ///
      (e(b_mi)[1,3]) (sqrt(e(V_mi)[3,3])) (e(b_mi)[1,1])
(sqrt(e(V_mi)[1,1])) ///
      (e(b_mi)[1,2]) (sqrt(e(V_mi)[2,2])) ///
      (0) (0) (0) (0) (0) (0) (0) (0) (0) (0) (0)

      *2. Missingness in confounder C
      drop _all
      quietly use sim1_DGM1000 if sampno==`i', clear
      quietly gen y=y1_b1_p1 if `phi' < 0.2
      quietly replace y=y1_b1_p6 if `phi' > 0.2 & `phi' < 0.7
```

```
      quietly replace y=y1_b1_1 if `phi' > 0.7
      gen x=x1_b
      if `tau' < 0.2 local alpha = 8
         else if `tau' > 0.2 & `tau' < 2 local alpha = 0.6
         else if `tau' > 2 & `tau' < 4 local alpha = 0.2
         else local alpha = 0.1

      quietly gen r_c3=rbinomial(1,invlogit(`tau'*(`alpha' + x)))
* Allowable range for p is 1e-8 to 1-1e-8, so extreme values of x
give missing values for r_c3
      quietly replace r_c3=0 if invlogit(`tau'*(`alpha' + x)) <
0.00000001
      gen c3miss=c3bin
      quietly replace c3miss=. if r_c3==0

      *CRA
      quietly regress y x c3miss
      *store values for -post-
      est store vals

      *Diagnostic tests
      *Tests for analysis model mis-specification (linear
regression)
      quietly regress y x c3miss
      *store values for -post-
      est store vals

      *Link test
      quietly linktest
      local link=(2 * ttail(r(df), abs(r(t))))
      * Shapiro-Wilks
      quietly swilk r
      local swilk=r(p)
      *Test for heteroskedasticity
      quietly estat hettest
      local het=r(p)
      * FP tests
      *first using default of dim(2)
      quietly predict r,resid
      quietly predict fit,xb
      quietly fp <fit>, scale: regress r <fit>
      local fp_d2=e(fp_compare)[1,4]
      *then using dim(1)
      drop fit_1 fit_2
      quietly fp <fit>, dim(1) scale: regress r <fit>
      local fp_d1=e(fp_compare)[1,4]
      *Grouped test
      xtile fit_q = fit, nq(5)
      quietly anova r fit_q
      local quint=Ftail(e(df_m),e(df_r),e(F))

      *Tests for imputation model mis-specification (logistic
regression)
      quietly logistic c3miss y x
      *Link test
      quietly linktest
```

```
      local link_lgt=(2 *  (1-normal(abs(r(t)))))
      *Hinkley test - using capture as does not always converge
      drop fit fit_1 fit_q
      quietly predict fit,xb
      capture {
      quietly fp <fit>, scale: logit c3miss <fit> y x
      *Compare to model without fit
      local hinkley=e(fp_compare)[1,4]
      }
      *If error, run rest of loop
      if _rc!=0 {
          local hinkley=.
            }
      *Hosmer-Lemeshow
      quietly estat gof, group(10)
      local HL=r(p)

      *Restore regression estimates and post all ests
      quietly est restore vals
      post `simloop' (`i') (`tau') (`phi') ("No") ("B1") ("X1")
("C") ("CRA") ///
      (_b[_cons]) (_se[_cons]) (_b[x]) (_se[x]) (_b[c3miss])
(_se[c3miss]) ///
      (0) (0) (`link') (`swilk') (`het') (`fp_d2') (`fp_d1')
(`quint') (`link_lgt') (`hinkley') (`HL')

      *MI linear imputation model
      *Note PMM not used because C is binary in scenario 1
      quietly mi set flong
      quietly mi register imputed c3miss
      quietly mi register regular y x
      quietly mi impute chained (logit) c3miss = y x, add(30)
      quietly mi estimate: regress y x c3miss
      post `simloop' (`i') (`tau') (`phi') ("No") ("B1") ("X1")
("C") ("MI") ///
      (e(b_mi)[1,3]) (sqrt(e(V_mi)[3,3])) (e(b_mi)[1,1])
(sqrt(e(V_mi)[1,1])) ///
      (e(b_mi)[1,2]) (sqrt(e(V_mi)[2,2])) ///
      (0) (0) (0) (0) (0) (0) (0) (0) (0) (0) (0)
}
}
}
postclose `simloop'
postclose `simseed'
```

*Section S12. Stata code for the real data analysis*

```
*Acupuncture data can be accessed here:
*https://www.ncbi.nlm.nih.gov/pmc/articles/PMC1489946/#S1
use acupuncture, clear

*CRA
regress pk5 age sex migraine chronicity pk1 group

* Test for model misspecification
quietly predict r,resid
quietly predict fit,xb
fp <fit>, scale: regress r <fit>

*MI linear imputation model
quietly mi set flong
quietly mi register imputed pk5
quietly mi register regular age sex migraine chronicity pk1 group
mi impute chained (regress) pk5=age sex migraine chronicity pk1
group, add(25)
mi estimate: regress pk5 age sex migraine chronicity pk1 group

*PMM
*Note, type 1 PMM can only be run in ice not mi impute
*First restore data i.e. before imputation
use acupuncture, clear
ice pk5 age sex migraine chronicity pk1 group, saving(tmp, replace)
m(25) match matchpool(5) uvisopts(matchtype(1))
use tmp, clear
quietly mi import ice, automatic
mi estimate: regress pk5 age sex migraine chronicity pk1 group

*Check functional form of each continuous variable
use acupuncture, clear
fp <age>, scale: regress pk5 <age> sex migraine chronicity pk1 group
*No evidence against linear form

fp <chronicity>, scale: regress pk5 age sex migraine <chronicity>
pk1 group
*No evidence against linear form

fp <pk1>: regress pk5 age sex migraine chronicity <pk1> group
*Suggests using squared version of pk1
gen pk1_sq=pk1^2

*Rerun mis-specification test on this model
regress pk5 age sex migraine chronicity pk1 pk1_sq group
drop r fit
quietly predict r,resid
quietly predict fit,xb
fp <fit>, scale: regress r <fit>

/* Finally, add squared term to the imputation model*/
quietly mi set flong
quietly mi register imputed pk5
```

```
quietly mi register regular age sex migraine chronicity pk1 group
pk1_sq
mi impute chained (regress) pk5=age sex migraine chronicity pk1
group pk1_sq, add(25)
mi estimate: regress pk5 age sex migraine chronicity pk1 group
```
